# Novel *CYP1B1-RMDN2* Alzheimer’s disease locus identified by genome-wide association analysis of cerebral tau deposition on PET

**DOI:** 10.1101/2023.02.27.23286048

**Authors:** Kwangsik Nho, Shannon L. Risacher, Liana Apostolova, Paula J. Bice, Jared Brosch, Rachael Deardorff, Kelley Faber, Martin R. Farlow, Tatiana Foroud, Sujuan Gao, Thea Rosewood, Jun Pyo Kim, Kelly Nudelman, Meichen Yu, Paul Aisen, Reisa Sperling, Basavaraj Hooli, Sergey Shcherbinin, Diana Svaldi, Clifford R. Jack, William J. Jagust, Susan Landau, Aparna Vasanthakumar, Jeffrey F. Waring, Vincent Doré, Simon M. Laws, Colin L. Masters, Tenielle Porter, Christopher C. Rowe, Victor L Villemagne, Logan Dumitrescu, Timothy J. Hohman, Julia B. Libby, Elizabeth Mormino, Rachel F. Buckley, Keith Johnson, Hyun-Sik Yang, Ronald C. Petersen, Vijay K. Ramanan, Prashanthi Vemuri, Ann D. Cohen, Kang-Hsien Fan, M. Ilyas Kamboh, Oscar L. Lopez, David A. Bennett, Muhammad Ali, Tammie Benzinger, Carlos Cruchaga, Diana Hobbs, Philip L. De Jager, Masashi Fujita, Vaishnavi Jadhav, Bruce T. Lamb, Andy P. Tsai, Isabel Castanho, Jonathan Mill, Michael W. Weiner, Alzheimer’s Disease Neuroimaging Initiative (ADNI), the Alzheimer’s Disease Neuroimaging Initiative – Department of Defense, the Anti-Amyloid Treatment in Asymptomatic Alzheimer’s Study (A4 Study), Australian Imaging, Biomarker & Lifestyle Study (AIBL), Andrew J. Saykin

## Abstract

Determining the genetic architecture of Alzheimer’s disease (AD) pathologies can enhance mechanistic understanding and inform precision medicine strategies. Here, we performed a genome-wide association study of cortical tau quantified by positron emission tomography in 3,136 participants from 12 independent studies. The *CYP1B1-RMDN2* locus was associated with tau deposition. The most significant signal was at rs2113389, which explained 4.3% of the variation in cortical tau, while *APOE4* rs429358 accounted for 3.6%. rs2113389 was associated with higher tau and faster cognitive decline. Additive effects, but no interactions, were observed between rs2113389 and diagnosis, *APOE4*, and Aβ positivity. *CYP1B1* expression was upregulated in AD. rs2113389 was associated with higher *CYP1B1* expression and methylation levels. Mouse model studies provided additional functional evidence for a relationship between *CYP1B1* and tau deposition but not Aβ. These results may provide insight into the genetic basis of cerebral tau and novel pathways for therapeutic development in AD.

---

Alzheimer’s disease (AD) is a common neurodegenerative disease featuring hallmark amyloid-beta (Aβ) plaques and neurofibrillary tau tangles.^1^ Aβ and tau measurements using positron emission tomography (PET) are common in research (i.e., amyloid/tau/neurodegeneration (A/T/N) classification criteria).^2^

Genetic factors conferring susceptibility to or protection from AD are important for identifying target biological pathways for drug development and personalized medicine.^3^ Large-scale genome-wide association studies (GWAS) using case-control designs have identified risk genes in immune, tau, Aβ, lipid, and other pathways.^4,5^ The strongest AD genetic risk locus is *APOE* (apolipoprotein E) ε4 (*APOE4*).^6^ Large case-control studies are often limited because participant neuropathology is unknown. Endophenotype studies of *in vivo* neuropathology complement case-control studies by testing genetic variants against disease pathology.^7^

Numerous studies have assessed genetic predictors of Aβ PET measures.^8-13^ However, most genetic studies of tau have utilized cerebrospinal fluid (CSF) tau measures due to non-availability of large tau PET datasets.^14^ One study investigated the association of [^18^F]flortaucipir PET with *BIN1*, finding an association between a known *BIN1* risk single nucleotide polymorphism (SNP; rs744373) and greater tau.^15^ Another performed GWAS on tau PET endophenotypes and identified two genetic loci (*PPP2R2B* and *IGF2BP3*). However, this study had limited statistical power due to the modest sample size (n=754) and did not include a replication sample.^16,17^

Here, we performed the largest GWAS of PET-based cortical tau to date (n=3,046). We included data from twelve independent cohorts. We also assessed the relationship of the top SNP with cognitive decline and additive and interaction effects with diagnosis, *APOE4* status, and Aβ positivity. We mapped topographic distribution of the top variant effect on voxel-wise tau deposition. We performed a gene-set enrichment analysis, assessed gene expression levels in human brain tissue and single-nucleus RNA-Seq data, mapped the expression of the top genes in the Allen Human Brain Atlas, and did methylation and expression quantitative trait loci (eQTL) analyses. Finally, we investigated expression levels of the top gene in tau and Aβ mouse models.^18-20^

## Results

### Genome-wide association analysis (GWAS)

Additive genetic models were tested for each SNP with transformed cortical tau covaried for for age, sex, population stratification, *APOE4* status, and diagnosis. We meta-analyzed GWAS results from seven cohorts in the discovery stage (n=1,446), the results of which are shown as quantile-quantile (**Figure 1A**) and Manhattan (**Figure 1B**) plots. No systematic *p*-value inflation was found (genomic inflation factor λ=1.025; **Figure 1A**). We identified a genome-wide significant association of cortical tau with a novel locus at 2p22.2 (**Figure 1B**), with two SNPs in the region reaching genome-wide significance (*p*-value ≤ 5×10^-8^). The strongest associated SNP for cortical tau is rs2113389, which was directly genotyped. The other SNP (rs918804) is in strong linkage disequilibrium (LD, r^2^ = 0.91 and D’ = 0.95) with rs2113389. The SNP (rs2113389) is located on 2p22.2 between *RMDN2* and *CYP1B1* and non-coding RNA, *CYP1B1- AS1* (**Figure 1C**). The minor allele T of rs2113389 (MAF=0.146) was associated with higher tau (Z score=5.68; *p*-value=1.37 x 10^-8^) and remained significant after including Aβ positivity as a covariate. We conducted a replication meta-analysis in five additional cohorts (n=1,600). The genome-wide significant SNPs (rs2113389 and rs918804) in the discovery stage were replicated with the same association direction (Z score=3.83, *p*-value=1.26 x 10^-4^; Z-score=-2.97, *p*- value=2.97×10^-3^, respectively; **Supplemental Figure 1**). We estimated the proportion of variance in cortical tau explained by these genetic variants using the Genome-wide Complex Trait Analysis (GCTA) tool^21^ and found that ∼4.3% of the variation in cortical tau in ADNI is explained by rs2113389 and *APOE4* SNP rs429328. In ADNI, rs2113389 was also associated with 1-year executive function decline after adjustment for age, sex, education, *APOE4* status, and baseline function (n=1,466; β=-0.053; *p*-value=0.014). Participants with the minor allele T of rs2113389 showed faster decline relative to non-carriers.

**Figure 1.**
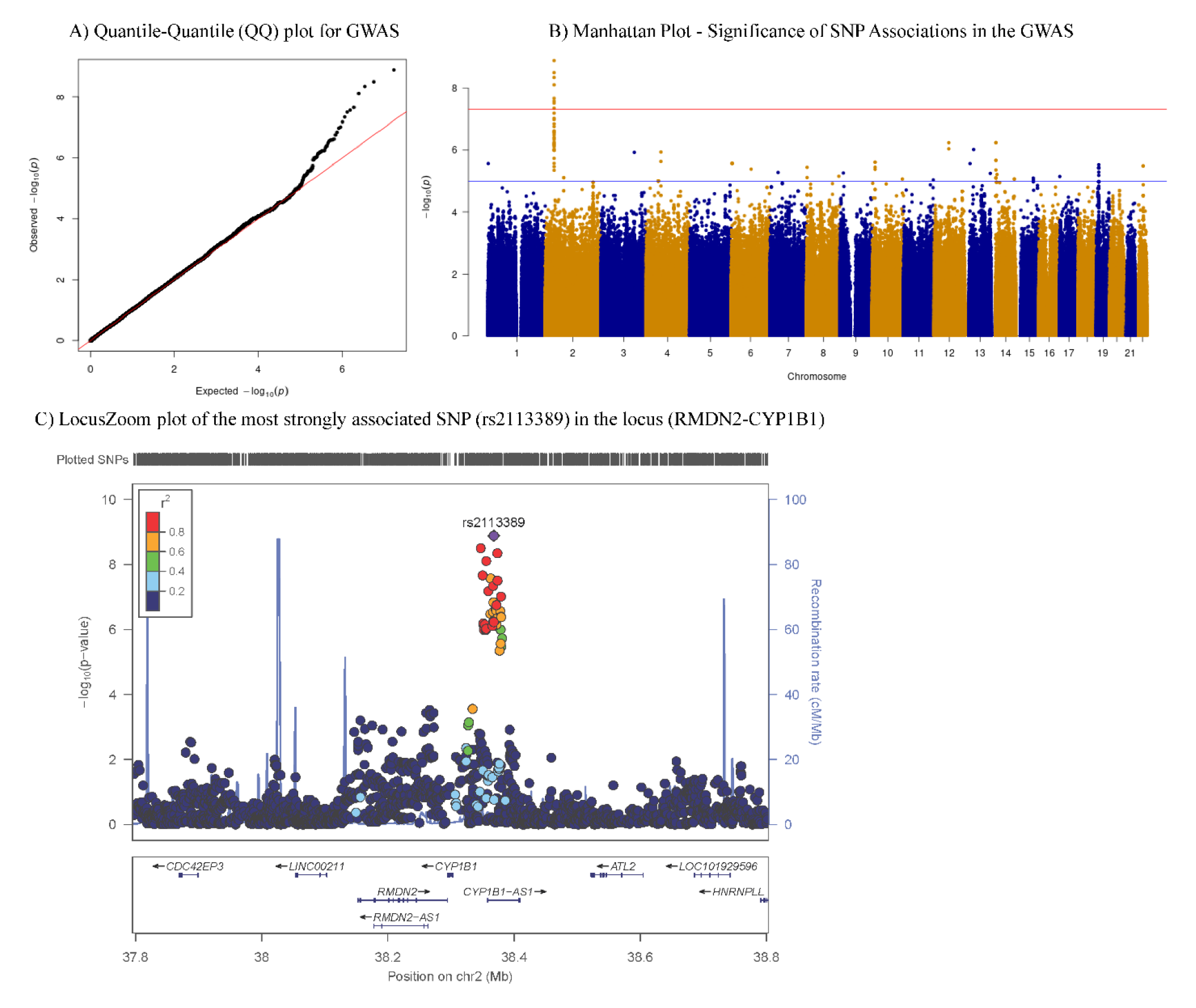
Results of Discovery GWAS for cortical tau deposition. Quantile-quantile (QQ) (A), Manhattan (B), and LocusZoom (C) plots of genome-wide association study (GWAS) results are shown. The genomic inflation factor is λ=1.025 In the Manhattan plot (B), the horizontal blue and red lines represent the -log_10_(10^−5^) and -log_10_(5.0 × 10^−8^) threshold levels, respectively. Two single nucleotide polymorphisms (SNPs) on chromosome 2 showed highly significant (<5.0 × 10^−8^) associations with cerebral tau deposition. The regional association plot (C) for the locus that passed genome-wide significance shows the region around the most significant SNP (rs2113389) at the *RMDN2-CYP1B1* locus. SNPs were plotted based on their GWAS −log_10_ *p*-values and genomic position. The red color scale of *r*^2^ values was used to label SNPs based on their degree of linkage disequilibrium with the most significant SNP. Recombination rates calculated from 1000 Genomes Project reference data are also displayed in a blue line corresponding to the right vertical axis. *Note: cerebral tau endophenotype measured as an inverse normal transformed variable of cortical tau SUVR*.

### Association of rs2113389 genotype with regional and global tau

**Figure 2** shows that both the additive (**Figure 2A&B**) and dominant models (**Figure 2C&D**) demonstrated higher MTL and cortical tau deposition in carriers of the minor allele (T) of rs2113389. Similar results were observed when stratified by sex (**Supplemental Figures 2 & 3**).

**Figure 2.**
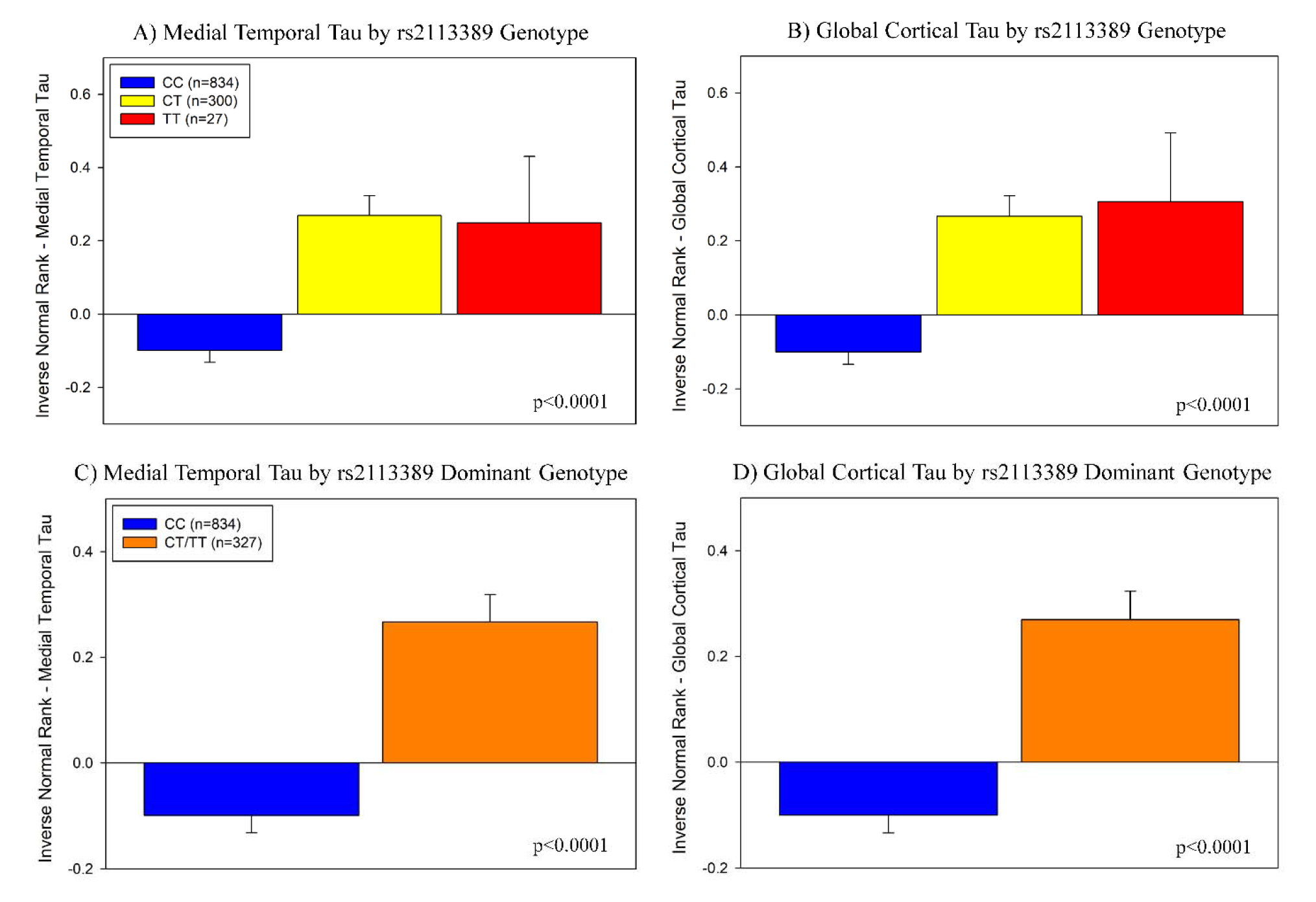
Association of the most significant SNP (rs2113389) at the *RMDN2-CYP1B1* locus with regional and global cortical tau burden. Using an additive model, the minor allele (T) of rs2113389 was associated with higher tau deposition across participants, with both rs2113389 CT and TT individuals showing significantly greater medial temporal lobe (A) and cortical (B) tau deposition than rs2113389 CC individuals. Similar results were seen using a dominant model. Specifically, individuals with one or more minor alleles of rs2113389 showed significantly greater tau deposition in the medial temporal lobe (C) and cortex (D) than rs2113389 CC individuals. All associations were significant at p<0.0001. *Note: tau measured as an inverse normal transformed variable of medial temporal and cortical tau SUVR*

### Interaction of rs2113389 genotype with variables of interest

Main effects of diagnosis and rs2113389 genotype were observed, with a higher MTL and cortical tau across diagnoses, but there was no interaction effect with rs2113389 dominant genotype (**Figure 3A&B**). The effect was similar in both males and females (**Supplemental Figure 4**). Main effects, but no interaction effect, for rs2113389 genotype and *APOE4* status were also observed (**Figure 3C&D**), with those positive for both *APOE4* and rs2113389 minor allele (T) showing the highest MTL and cortical tau. The sex-stratified analysis showed similar results in both males and females (**Supplemental Figure 5**). Finally, main effects of Aβ positivity and rs2113389 genotype, but no interaction effect, on MTL and cortical tau were observed (**Figure 3E&F**), with Aβ+ carriers of the rs2113389 minor allele (T) showing the highest tau. In the sex-stratified analysis, males and females showed similar results (**Supplemental Figure 6A&B**), except for an interaction effect of Aβ positivity and rs2113389 genotype on MTL tau deposition in females (**Supplemental Figure 6C**) and a trend for cortical tau (**Supplemental Figure 6D**).

**Figure 3.**
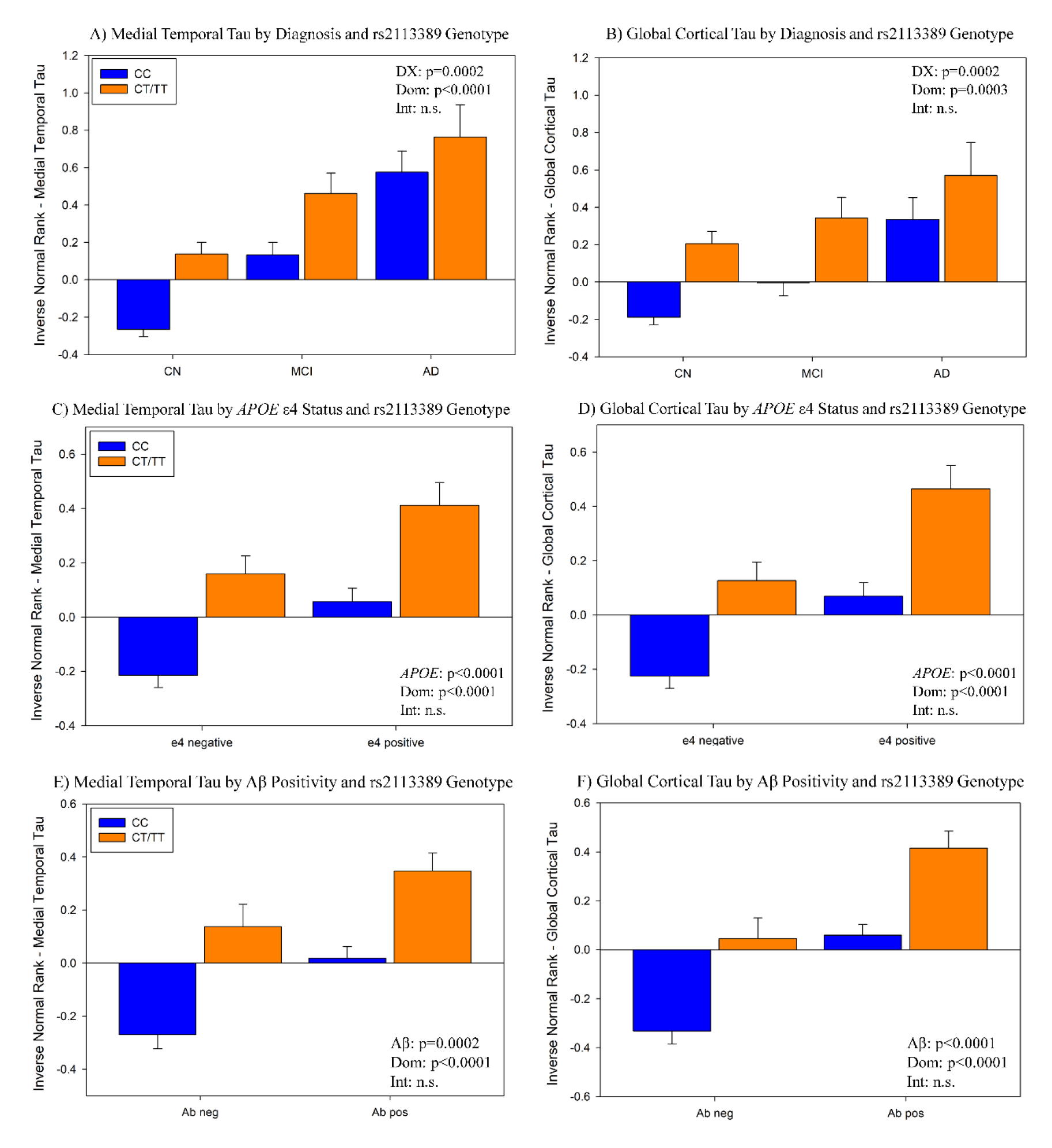
Interaction effect of the most significant SNP (rs2113389) at the *RMDN2-CYP1B1* locus with diagnosis (A) and (B), *APOE4* carrier status (C) and (D), and Aβ positivity (E) and (F) on regional and cortical tau deposition. Both diagnosis and rs2113389 dominant genotype were significantly associated with medial temporal (A) and cortical (B) tau deposition (all p<0.001), such that AD participants who carry at least one minor allele (T) of rs2113389 have the highest tau level relative to all other diagnoses and CC rs2113389 individuals. AD=Alzheimer’s disease; CN=cognitively normal; DX=diagnosis; Dom=rs2113389 dominant genotype (CC vs. CT/TT); Int.=interaction; MCI=mild cognitive impairment; n.s.=not significant. *APOE4* carrier status and rs2113389 dominant genotype are significantly associated with medial temporal (C) and cortical (D) tau deposition (all p<0.0001). The highest tau level is observed in carriers of both at least one *APOE4* allele and minor allele (T) at rs2113389, relative to either CC rs2113389 individuals or *APOE4* negative individuals. APOE=apolipoprotein E. Significant effects of both Aβ positivity and rs2113389 dominant genotype on medial temporal (E) and cortical (F) tau deposition are observed (all p<0.001), with Aβ positive individuals carrying at least one minor allele (T) at rs2113389 showing the highest level of tau deposition relative to all other groups (Aβ negative individuals, rs2113389 CC individuals). Aβ=amyloid-beta. *Note: tau measured as an inverse normal transformed variable of medial temporal and cortical tau SUVR*

### Voxel-wise association of rs2113389 genotype with tau

A voxel-wise analysis of the effect of rs2113389 (voxel-wise p<0.05 (FWE corrected), minimum cluster size (k)=100 voxels; **Figure 4** and **Supplemental Figure 7**) was completed to evaluate the topographic pattern of the association. In the dominant model, individuals carrying at least one minor allele at rs2113389 (CT or TT; n=327) demonstrated greater tau throughout the temporal lobe, parietal lobe, and inferior frontal lobe than rs2113389 CC individuals (n=834; **Figure 4A**). Beta-value maps supported the statistical map, showing widespread areas where rs2113389-T carriers show higher tau than non-carriers (**Figure 4B**). Using an additive model, rs2113389 CT individuals (n=300) showed higher tau than CC individuals (n=834) in the temporal, lateral parietal, and frontal lobes (**Supplemental Figure 7A**), while rs2113389 TT (n=27) showed a more focal region of higher frontal tau relative to rs2113389 CC individuals (**Supplemental Figure 7B**). Beta-value maps revealed interesting patterns, with rs2113389 CT individuals showing higher temporal and parietal tau relative to rs2113389 CC individuals (**Supplemental Figure 7C**). Alternatively, rs2113389 TT individuals showed widespread higher tau relative to rs2113389 CC individuals, especially in the frontal lobe (**Supplemental Figure 7D**). Finally, despite not reaching statistical significance likely due to power issues, the beta-values map shows that rs2113389 TT homozygotes show higher frontal tau than rs2113389 CT heterozygotes (**Supplemental Figure 7E**). These findings may suggest later Braak stages are more likely in TT homozygotes or that TT homozygotes have a cortical rather than limbic pattern relative to CT heterozygotes and CC homozygotes.

**Figure 4.**
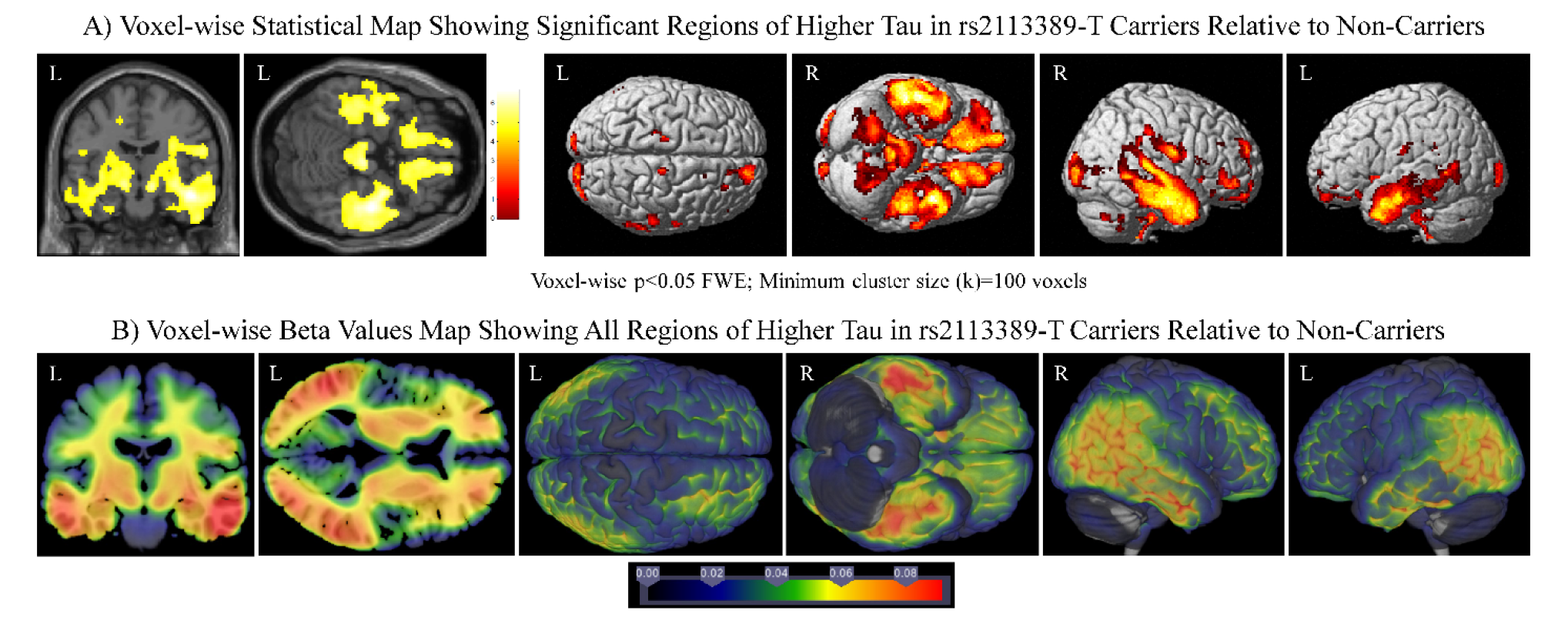
Voxel-wise analysis and visualization of the effect of rs2113389 dominant genotype on tau deposition. (A) Widespread regions of association between rs2113389 dominant genotype and tau deposition are observed in the inferior frontal, parietal, and medial and lateral temporal lobes, such that those with one or more minor alleles (T) at rs2113389 show greater tau deposition than CC rs2113389 individuals. Images are displayed at a voxel-wise threshold of p<0.05 with family-wise error correction for multiple comparisons and a minimum cluster size (k)=100 voxels. (B) Beta-value maps show widespread regions of higher tau deposition in rs2113389-T carriers relative to non-carriers. Specifically, temporal, parietal, and frontal lobe tau is greater in minor allele carriers than non-carriers.

### Pathway analysis

When gene ontology (GO) terms were considered, 480 gene-sets were significant after correction for multiple testing. GO for cell-cell adhesion was the most significant pathway identified (**Table 1A**). GO terms for MHC protein complex, postsynaptic density, regulation of synaptic transmission, and calcium ion transport were also significant. For the Kyoto Encyclopedia of Genes and Genomes (KEGG) pathway, 44 gene-sets were significant, including cell adhesion molecules, calcium signaling pathways, and axon guidance (**Table 1B**).

**Table 1.**
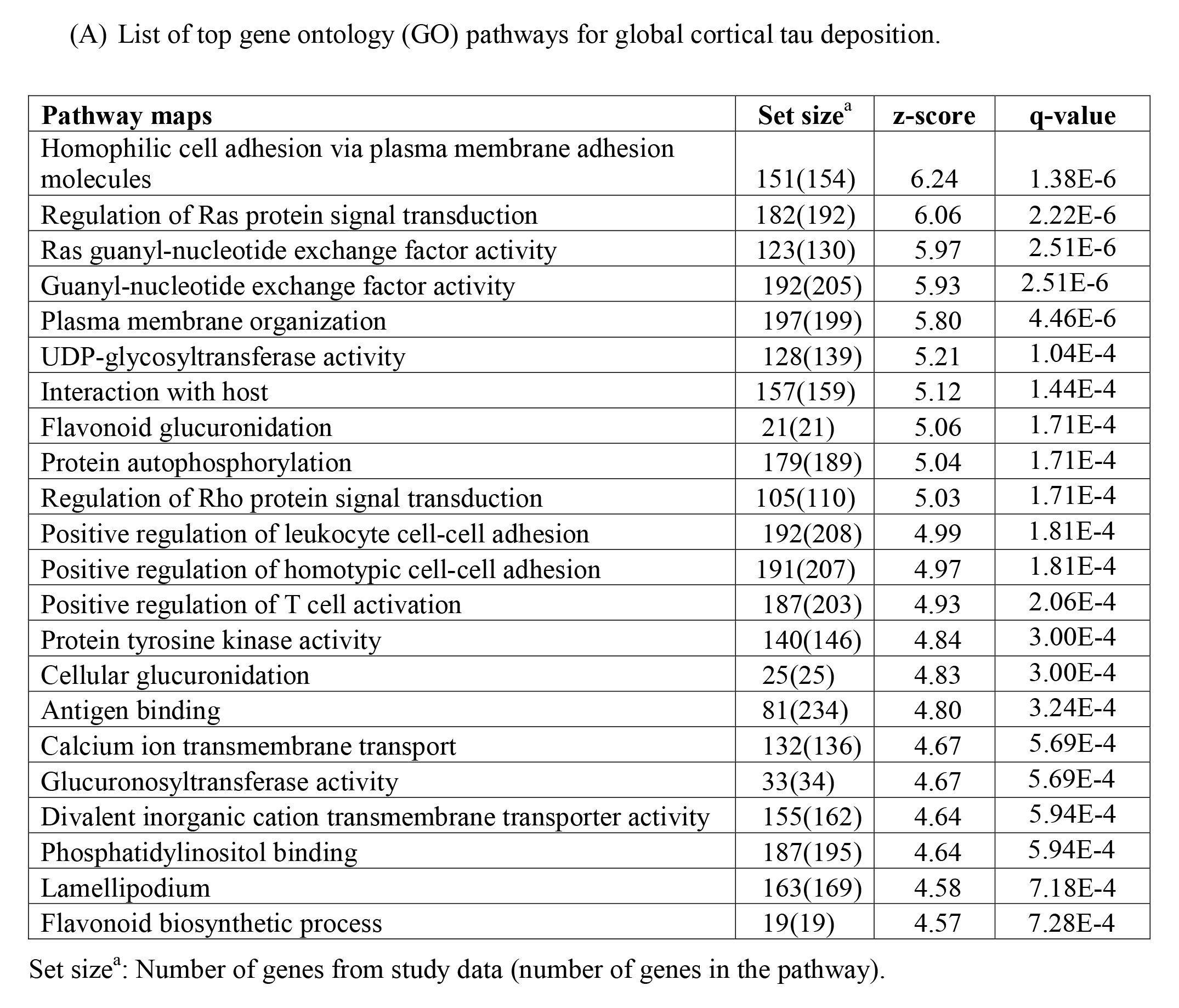
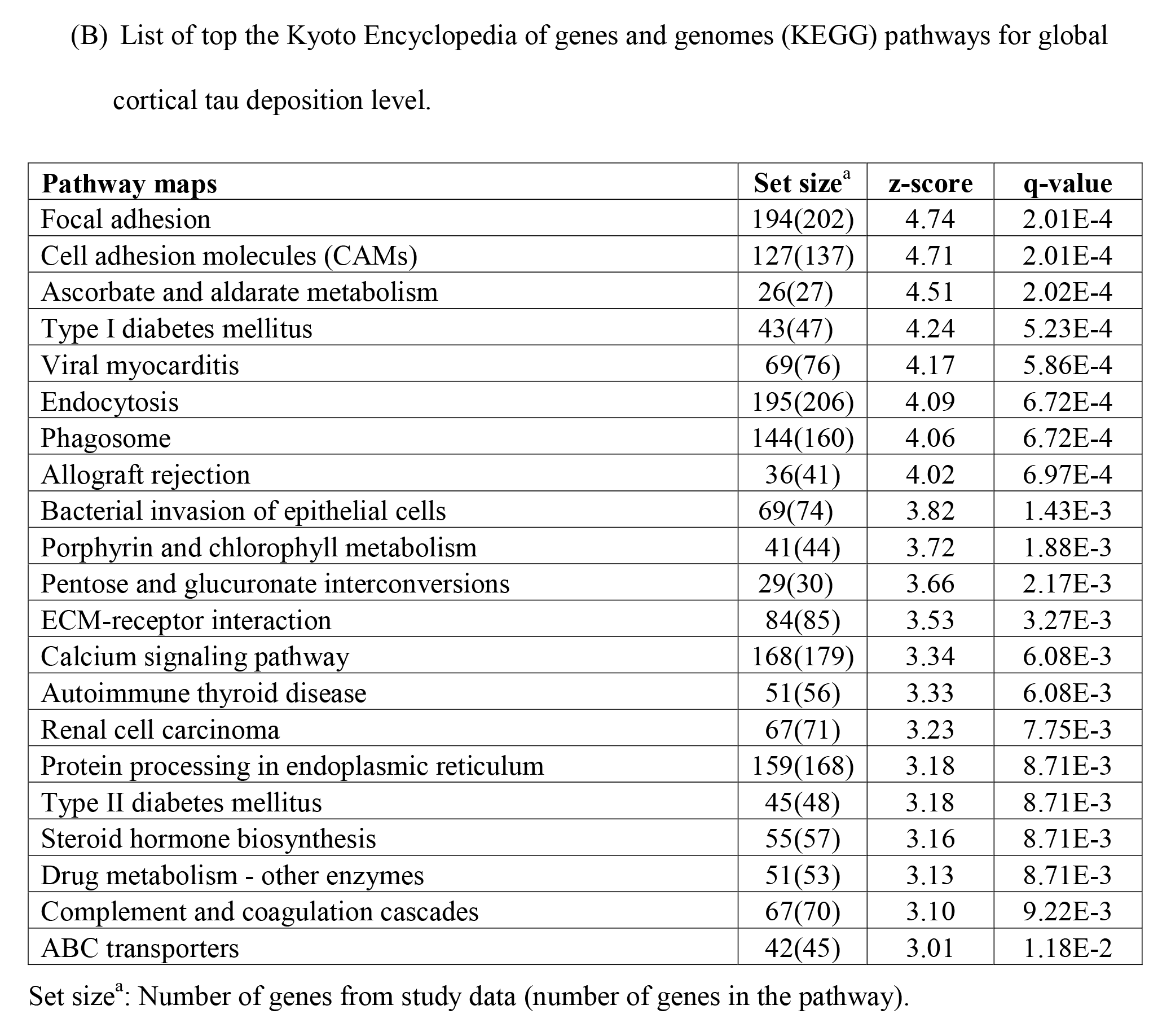
Gene set enrichment analysis of global cortical tau deposition.

### Gene expression analysis and eQTL analysis

Our genome-wide gene-based association analysis identified two protein coding genes (*CYP1B1* (corrected *p*-value=0.040), *RMDN2* (corrected *p*-value=0.040)), and one non-coding RNA (*CYP1B1-AS1* (corrected *p*-value=0.040)) as associated with cortical tau after multiple testing adjustment. Then, our Allen Human Brain Atlas visualization showed that *CYP1B1* was highly expressed across the whole brain, especially in the insula, orbitofrontal cortex, temporal lobe, and medial temporal lobe. *RMDN2* was also highly expressed across the whole brain, especially in the temporal lobe, visual cortex, frontal (medial orbitofrontal cortex) and posterior (precuneus and isthmus cingulate cortex) default mode network regions, and sensorimotor cortex (**Supplemental Figure 8**). Bulk RNA-Seq data from 1,917 samples preprocessed in AMP-AD was evaluated for these genes. Differential expression of *RMDN2* was seen in the parahippocampal gyrus (*p*-value=0.004; **Figure 5A**), with down-regulation in AD. *CYP1B1* demonstrated differential expression in the temporal cortex (*p*-value=0.001; **Figure 5B**), with upregulation in AD. We also investigated whether the identified SNPs were associated with expression levels of *CYP1B1* and *RMDN2* (eQTL). The most significantly associated SNP, rs2113389, was associated with *CYP1B1* expression levels in the temporal cortex, but not with *RMDN2* expression. Specifically, the rs2113389 T-allele was associated with higher temporal *CYP1B1* expression (β=0.25; *p*-value=0.02; **Figure 5C**). Finally, the rs2113389 T-allele was associated with higher *CYP1B1* expression levels in blood from the eQTLGen consortium database (n=31,684; Z score=24.93; *p*-value= 3.6 x 10^-137^).

**Figure 5.**
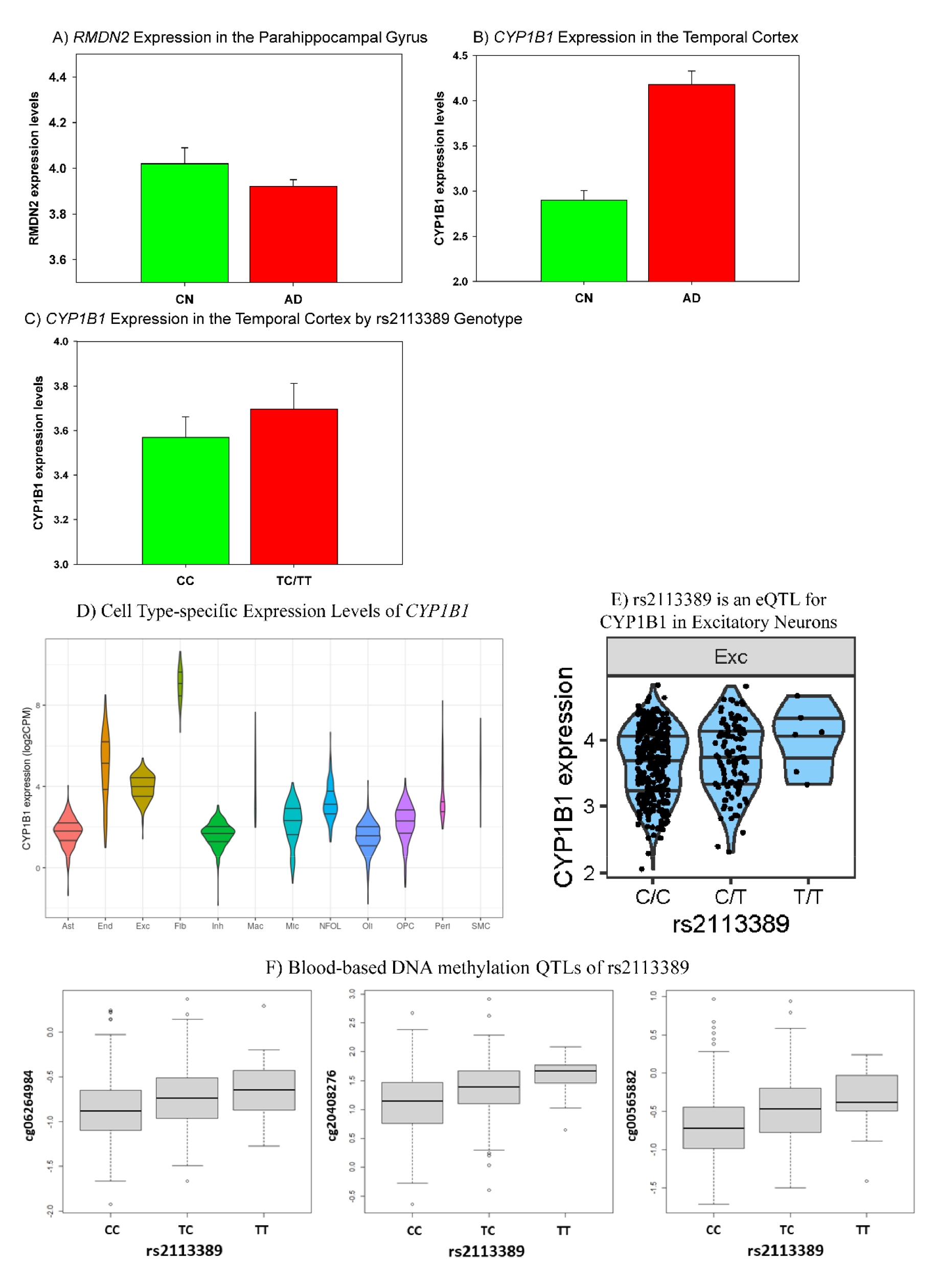
Gene expression analysis of *RMDN2* and *CYP1B1* and expression quantitative trait locus (eQTL) and DNA methylation QTL (meQTL) analysis of rs2113389. Patients with AD showed downregulated expression of *RMDN2* in the parahippocampal gyri (A) and upregulated expression of *CYP1B1* in the temporal cortex (B) relative to CN individuals using brain tissue-based RNA-Seq data from the AMP-AD project. (C) In an eQTL analysis, the identified SNP (rs2113389) was shown to be associated with *CYP1B1* expression levels in the temporal cortex, with carriers of the minor allele showing upregulated *CYP1B1* expression relative to individuals with the rs2113389 CC genotype. AD=Alzheimer’s disease; CN=cognitively normal. Cell type-specific expression levels (D) and eQTL in the excitatory neuron (E) of *CYP1B1* gene are shown. In (D), the x-axis is cell types in ROSMAP DLPFC single-nucleus RNA-Seq data. The y-axis is the log_2_ of counts per million mapped reads (CPM) of *CYP1B1* gene. Bars show the 25%, 50%, and 75% quartiles, respectively. Expression levels were computed at the donor level, by aggregating cells from the same donor. Rare cell types were observed only in a small fraction of donors. To reflect this, areas of violin plots are scaled proportionally to the number of donors. Fibroblasts (Fib) had the highest expression of *CYP1B1* gene. Among major cell types, excitatory neurons (Exc) had the highest expression. In (E), the minor allele (T) of rs2113389 was associated with higher cell type-specific *CYP1B1* expression levels in the excitatory neuron (*p*-value= 0.035). DNA methylation QTL analysis (F; *cis*- meQTL) of rs2113389 with CpGs in *CYP1B1* measured in blood samples from 634 ADNI participants identified three CpGs, located in the *CYP1B1* gene body region, as significantly associated with rs2113389 (*p*-value < 1 x 10^-5^). The minor allele (T) of rs2113389 was associated with higher expression levels of the CpGs.

### Cell type-specific expression and eQTL analysis of CYP1B1

Single-cell expression of *CYP1B1* in ROSMAP single-nucleus RNA-Seq data from the dorsolateral prefrontal cortex showed that fibroblasts (Fib) had the highest expression of the *CYP1B1* gene across all cell types. Among the eight major brain cell types, excitatory neurons (Exc) had the highest expression of *CYP1B1* (**Figure 5D**). Finally, eQTL analysis of cell type specific *CYP1B1* expression in excitatory neurons showed that the rs2113389 T-allele was associated with higher cell type-specific *CYP1B1* expression levels (*p*-value= 0.035; **Figure 5E**).

### Blood-based DNA methylation QTLs of rs2113389

The DNA methylation QTL analysis (meQTL) of rs2113389 with CpGs in *CYP1B1* measured in blood identified three CpGs located in the *CYP1B1* gene body region associated with rs2113389 (*p*-value < 1 x 10^-5^; **Figure 5F**). The rs2113389 T-allele was associated with higher CpG expression levels.

### Cyp1b1 expression and expression changes in the brain of AD mice

*Cyp1b1* expression was increased in the cortex of 6-month-old hTAU mice, consistent with our findings in humans (*p*-value=0.038; **Figure 6A**). *Cyp1b1* expression also significantly changed with time (genotype*age) in rTg4510 mice (FDR corrected *p*-value=0.040) but not J20 mice relative to wild-type mice (**Figure 6B** and **Figure 6C**). *Cyp1b1* differential expression over time in the TG rTg4510 mice was associated with entorhinal cortex tau pathology (FDR-corrected *p*- value=0.002; **Figure 6B** and **Figure 6C**).

**Figure 6.**
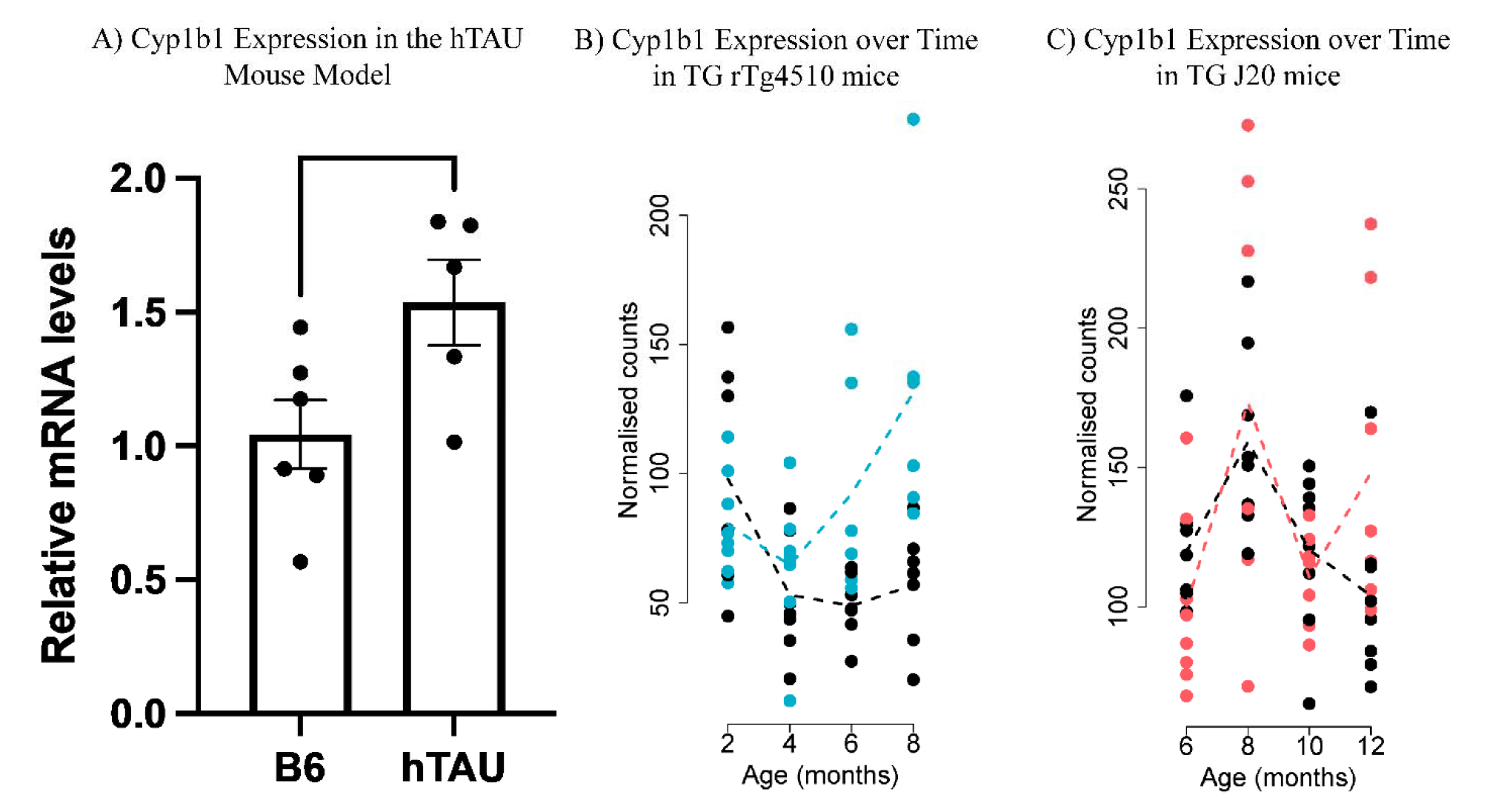
*Cyp1b1* expression and expression changes in the brain of tau (hTAU, rTg4510) and amyloid (J20) mice. (A) *Cyp1b1* expression was increased in the hTAU mouse model expressing six isoforms of human tau, consistent with our findings in human LOAD (*p*-value=0.038). In hTAU mice, *Cyp1b1* expression was increased in the cortex of 6-month-old mice. (B) *Cyp1b1* expression significantly changed with time (genotype*age) in TG rTg4510 mice, i.e., *Cyp1b1* is associated with disease progression in the rTg4510 model. *Cyb1b1* is also specifically associated with tau pathology progression. (C) *Cyp1b1* expression did not change with time (genotype*age) in J20 mice, i.e., *Cyp1b1* is not associated with disease progression in the J20 model. In addition, *Cyb1b1* is not associated with amyloid pathology progression.

## Discussion

We performed a genome-wide association analysis (GWAS) of cortical tau PET and identified and replicated a novel SNP at the *CYP1B1*-*RMDN2* locus at 2p22.2. The most significant SNP at the locus originated from rs2113389, with the minor allele (T) of rs2113389 associated with higher MTL and cortical tau. Higher tau levels in rs2113389 T-allele carriers were observed across diagnoses. An additive effect of the T-allele with *APOE4* status was also observed, with minor T-allele carriers and *APOE*4 carriers having the highest tau levels. Similar findings were seen with Aβ positivity, such that Aβ+ carriers of the rs2113389 T-allele had the highest tau level. In sex-stratified analyses, generally similar results were observed, except for an interaction of Aβ positivity and rs2113389 genotype on MTL tau in females. Overall, these results provide converging evidence that the minor allele (T) of rs2113389 is a risk variant for high tau. Voxel-wise whole brain analysis confirmed that the rs2113389 T-allele was associated with tau in AD-related cortical regions. These findings also support a previous GWAS of CSF tau, where rs1478361, which is in strong LD with rs2113389 (r^2^ = 0.96 and D’ = 1.00), was associated with CSF total tau levels (n=3,076; β=0.0176; *p*-value=0.0295).^14^

The two protein coding genes at the locus identified in this analysis (*CYP1B1* and *RMDN2*) are highly expressed in the brain in the frontal and temporal lobes for *CYP1B1* and in throughout the cortex for *RMDN2*. Of note, the regions showing higher expression levels overlap with the typical patterns of tau deposition. The finding suggests that there exists a spatial relationship between regional expression levels of the two genes and tau deposition patterns, supporting the selective vulnerability hypothesis of tau pathology. *RMDN2* (Regulator of Microtubule Dynamics 2) is down-regulated in the parahippocampal gyrus in AD, while *CYP1B1* (Cytochrome P450 Family 1 Subfamily B Member 1) is up-regulated in the temporal cortex in AD. The minor allele of rs2113389 is also associated with higher temporal cortex *CYP1B1* expression levels. Fibroblasts and excitatory neurons had the highest expression levels of the *CYP1B1* gene in the brain, and in excitatory neurons, the minor allele of rs2113389 was associated with higher expression levels of the *CYP1B1* gene. Blood-based DNA methylation analysis also supported the impact of the rs2113389 on CpGs within the *CYP1B1* gene, with the minor allele of rs2113389 associated with higher CpG expression. Finally, *Cyp1b1* expression was higher in the cortex of 6-month-old hTAU mice relative to controls. In addition, in the longitudinal analysis, *Cyp1b1* expression changed with aging in rTg4510 mice but not J20 mice, which suggests that *Cyp1b1* expression was associated with progression of tau but not amyloid pathology.

*CYP1B1* is of particular interest as the eQTL analysis shows altered temporal lobe expression in AD patients, and rs2113389 genotype is linked to the amount of temporal lobe *CYP1B1* expression. *CYP1B1* is a member of the cytochrome p450 enzyme family (CYP). CYP is present and active in the brain and expressed in a region- and cell-specific manner, including in the blood-brain barrier.^22-24^ CYP is responsible for oxidative metabolism of exogenous and endogenous substrates, potentially having both neuroprotective and pathologic roles.^23^ CYP is also involved in modulating blood flow, metabolism of fatty acids, cholesterol, and neurotransmitters, and mobilization of intracellular calcium,^25-28^ suggesting multiple potential roles in AD. Previously, genetic variants in CYP genes have been associated with neurodegenerative diseases, including AD,^29,30^ as well as AD pathophysiology (Aβ and tau).^25,31-33^ *CYP1B1* regulates endogenous pathways involved in metabolism of drugs and synthesis of cholesterols, steroids, and other lipids.^34^ While several cytochrome P450 family genes such as *CYP2C19* have been implicated in AD, *CYP1B1* has not previously been directly implicated in AD.^29,30,33^ However, *CYP1B1* may have multiple potential roles related to AD-related tau pathology and has been shown to be a regulator of oxidative stress, which in turn promotes angiogenesis.^35,36^ *CYP1B1* also promotes angiogenesis by suppressing NF-kB activity, which is also implicated in inflammation.^37^ Previous studies suggested that *CYP1B1* inhibition reduced oxidative stress and metabolized cell products that modulate intracellular oxidative stress; however, a lack of *CYP1B1* leads to increased intracellular oxidative stress in the endothelium.^38-40^ *CYP1B1* may play an important role in high fat diet-associated learning and memory deficits and oxidative damage.^40^ Increased brain oxidative stress causes damage to cell function with aging and is an important pathogenic factor in AD, contributing to tau phosphorylation and the formation of neurofibrillary tangles.^41-43^ Functional studies for *RMDN2* are limited, only showing that it encodes a protein important for regulating microtubule dynamics.

Pathway-based analysis identified enrichment in pathways related to the MHC, postsynaptic membrane, postsynaptic density, synapse organization, and calcium channel activity. MHC proteins and signaling have been implicated in large-scale AD genetic associations,^4,44,45^ along with associations with specific MHC alleles.^46^ Microglial activation via MHC class II signaling is increased in regions of phosphorylated tau.^47^ Dysfunctional synaptic connections are involved early in AD-related cognitive impairment,^48^ and tau deposition may induce synaptic impairment and learning deficits.^49,50^ Studies also suggest a role for tau at dendritic spines in affecting the trafficking of postsynaptic receptors.^51,52^ Finally, the “Calcium Hypothesis” suggests that Ca^2+^ signaling and homeostasis are implicated in AD pathology.^53^ Calcium signaling controls a variety of pathways, including activation of calpain, which has been shown to precede tau phosphorylation.^54,55^ Treatments targeting calcium channels are potential pathways for novel therapeutics for neurodegenerative diseases.^55^

There are some notable limitations, as studies are primarily observational and composed only of cohorts of European ancestry. Multiethnic studies are important, and to be generalizable to other populations, our findings require replication using large community studies or international collaborations. Although we performed the largest GWAS of tau PET to date, our meta-analysis had limited statistical power due to the moderate sample size for genetic association. Additional independent large cohorts with tau PET and GWAS data will enable validation studies.

In summary, GWAS of tau PET identified novel genetic variants in a locus (*CYP1B1*-*RMDN2*) that influences MTL and cortical tau. The mechanistic significance of this locus was supported by a range of independent functional genomic observations in humans and model systems. Taken together, these results can inform future biomarker and therapeutic development.

## Methods

### Participants

Participants from the Alzheimer’s Disease Neuroimaging Initiative (ADNI; http://adni.loni.usc.edu), ADNI-Department of Defense (ADNI-DoD), Indiana Memory and Aging Study (IMAS) of the Indiana ADRC, Avid A05 clinical trial (A05), Anti-Amyloid Treatment in Asymptomatic Alzheimer’s (A4) and Longitudinal Evaluation of Amyloid Risk and Neurodegeneration (LEARN) studies, Harvard Aging Brain Study (HABS), University of Pittsburgh Alzheimer’s Disease Research Center (UPitt ADRC), Mayo Clinic Study of Aging (MCSA), Memory and Aging Project (MAP) at the Knight Alzheimer’s Disease Research Center (Knight-ADRC), the Australian Imaging, Biomarker and Lifestyle Study (AIBL), and the Berkeley Aging Cohort Study (BACS) were included. The discovery sample included ADNI, ADNI-DoD, IMAS, A05, A4, HABS, UPitt ADRC. The replication sample included MCSA, MAP-Knight ADRC, AIBL, and BACS. *Post-hoc* analyses of interactions with diagnosis, *APOE4* status, and Aβ positivity, as well as voxel-wise analyses were performed in 1,161 individuals from ADNI, ADNI-DoD, IMAS, A05, A4, and LEARN. Informed consent was obtained for all participants, and studies were approved by the relevant institutional review boards. Descriptions of all cohorts, as well as demographics and cognitive performance, are found in the Supplemental information (**Supplemental Tables 1-12**).

### Genotyping and imputation

Participants were genotyped using several genotyping platforms. Un-genotyped SNPs were imputed separately in each cohort using the Haplotype Reference Consortium (HRC) data as a reference panel.^56^ Before imputation, standard sample and SNP quality control (QC) procedures were performed, as described previously.^57^ Furthermore, only non-Hispanic participants of European ancestry by multidimensional scaling analysis were selected.^58^ Imputation and QC procedures were performed as described previously.^59^

### Statistical analysis

#### Genome-wide association analysis (GWAS)

Cortical tau deposition followed a normal distribution after a rank-based inverse normal transformation. Using imputed genotypes, a GWAS of cortical tau was performed using a linear regression model with age, sex, two principal component (PC) factors from population stratification, *APOE4* status, and diagnosis as covariates using PLINK.^60^ A conservative threshold for genome-wide significant association (*p*<5 × 10^−8^) was employed.^61^ QQman was used to generate Manhattan and Q-Q plots, and LocusZoom was used to obtain regional association plots for selected loci.^62^

#### Gene-set enrichment analysis

Gene-set enrichment analysis was performed using GWAS summary statistics to identify pathways and functional gene sets associated with cortical tau deposition using the GSA-SNP software,^63^ as described in the Supplemental information. Enriched pathways with cortical tau levels were defined as those with FDR-corrected *p-* value<0.05.

#### Gene-based association analysis

Genome-wide gene-based association analysis was performed using GWAS p-values and the KGG software as described previously^64,65^ and in the Supplemental information.

#### Interaction with diagnosis, *APOE* genotype, and Aβ positivity

The effect of the top identified SNP (rs2113389 – dominant model) and its interaction with diagnosis, *APOE4* status, and Aβ positivity, on global and medial temporal lobe (MTL) tau, which is a primary location of early tau in AD, was assessed as described in the Supplemental information. Differential effects by sex were also evaluated using stratified analysis (methods in Supplemental information).

#### Detailed whole-brain imaging analysis

Tau PET SUVR images (n=1,161) were used in a voxel-wise statistical analysis of the effect of the top identified SNP on tau using SPM12 (www.fil.ion.ucl.ac.uk/spm/) in a *post-hoc* analysis (described in the Supplemental information). A voxel-wise threshold of p<0.05 with family-wise error (FWE) adjustment for multiple comparisons was used.

#### AMP-AD bulk RNA-Seq data in the post□mortem human brain

Pre-processed bulk RNA-Seq data from 1,917 samples were downloaded from Synapse (https://www.synapse.org/#!Synapse:syn17115987) of the AMP-AD Consortium^66-70^ and analyzed as discussed in the Supplemental information. Procedures for sample collection, post-mortem sample descriptions, tissue and RNA preparation methods, library preparation and sequencing methods, and sample quality controls were previously described.^70^ Finally, the eQTLGen^71^ consortium database (n=31,684) was used for eQTL of rs2113389 with *CYP1B1* expression in blood.

#### Single-nucleus RNA-Seq (snRNA-Seq) preprocessing and analysis

Frozen brain tissue specimens (n=479) from the dorsolateral prefrontal cortex were obtained in the Religious Orders Study/Memory and Aging Project (ROSMAP) cohort^72^ and processed as described in the Supplemental information. Raw data is available through the AD Knowledge Portal (https://www.synapse.org/#!Synapse:syn31512863). Fifty-five samples were excluded for quality control issues (see Supplementary information).

#### Allen Human Brain Atlas data and analysis

Regional gene expression profiles for *CYP1B1* and *RMDN2* were derived from brain-wide microarray-based transcriptome data from the Allen Human Brain Atlas.^73,74^ See Supplementary information for detailed methods.

#### ADNI DNA methylation data

In ADNI, Illumina EPIC chips (Illumina, Inc., San Diego, CA, USA) were used to profile DNA methylation in 1,920 blood or buffy coats samples including 200 duplicate samples according to the Illumina protocols. A detailed protocol has been published previously^77-79^ and briefly described in the Supplemental information. We performed methylation quantitative trait loci (meQTLs) of the top SNP discovered with CpGs in that gene (n=634) using multivariate linear models adjusted for age, sex, cell composition changes, and DNA storage/source.

#### AD pathology mouse model analysis

<u>hTau mouse model</u>: Generation of the hTAU mice, as well as brain extraction and tissue processing, was described previously^18,19,80,81^ and in the Supplemental information. Student’s t test was performed for qPCR results comparing C57BL/6J (B6; wild type) and hTAU mice. <u>rTg4510 and J20 mouse model</u>: Transgenic mice harboring human tau (rTg4510) and amyloid precursor protein (J20) mutations were used to investigate if gene expression changes of the top identified gene was associated with AD pathology.^20^ The rTg4510 and J20 mouse models and experimental models and methods were described previously,^20,82-85^ and are briefly summarized, along with statistical methods used, in the Supplemental information.

## Data Availability

All data are either available online at adni.loni.usc.edu https://ida.loni.usc.edu/collaboration/access/appLicense.jsp;jsessionid=2CC9FA6681E2924181B892706F917FEA or are available upon reasonable request from the authors.

https://adni.loni.usc.edu

https://ida.loni.usc.edu/collaboration/access/appLicense.jsp;jsessionid=2CC9FA6681E2924181B892706F917FEA

https://www.synapse.org/#!Synapse:syn17115987

https://www.synapse.org/#!Synapse:syn31512863

## Abbreviations

AD: Alzheimer’s disease
GWAS: genome-wide association study
SNP: single nucleotide polymorphism
CSF: cerebrospinal fluid
PET: positron emission tomography
APOE: apolipoprotein E
ADNI: Alzheimer’s Disease Neuroimaging Initiative
ADNI-DoD: ADNI-Department of Defense
IMAS: Indiana Memory and Aging Study
eQTL: expression quantitative trait loci
SUVR: standard uptake value ratio
FWHM: Full Width at Half Maximum
ROI: region of interest
HRC: Haplotype Reference Consortium
MAF: minor allele frequency
LASSO: Least Absolute Shrinkage and Selection Operator
CEU: Utah residents with Northern and Western European ancestry from the CEPH collection
TSI: Toscani in Italia
MDS: multidimensional scaling
CN: Cognitively normal older adults
MCI: Mild cognitive impairment
RMDN2: Regulator of Microtubule Dynamics 2
CYP1B1: Cytochrome P450 Family 1 Subfamily B Member 1

## Acknowledgements

Data collection and sharing for this project was funded by the Alzheimer’s Disease Neuroimaging Initiative (ADNI) (National Institutes of Health Grant U01AG024904) and DOD ADNI (Department of Defense award number W81XWH-12-2-0012). ADNI is funded by the National Institute on Aging, the National Institute of Biomedical Imaging and Bioengineering,and through generous contributions from the following: AbbVie, Alzheimer’s Association; Alzheimer’s Drug Discovery Foundation; Araclon Biotech; BioClinica, Inc.; Biogen; Bristol-Myers Squibb Company; CereSpir, Inc.; Cogstate; Eisai Inc.; Elan Pharmaceuticals, Inc.; Eli Lilly and Company; EuroImmun; F. Hoffmann-La Roche Ltd and its affiliated company Genentech, Inc.; Fujirebio; GE Healthcare; IXICO Ltd.; Janssen Alzheimer Immunotherapy Research & Development, LLC.; Johnson & Johnson Pharmaceutical Research & Development LLC.; Lumosity; Lundbeck; Merck & Co., Inc.; Meso Scale Diagnostics, LLC.; NeuroRx Research; Neurotrack Technologies; Novartis Pharmaceuticals Corporation; Pfizer Inc.; Piramal Imaging; Servier; Takeda Pharmaceutical Company; and Transition Therapeutics. The Canadian Institutes of Health Research is providing funds to support ADNI clinical sites in Canada. Private sector contributions are facilitated by the Foundation for the National Institutes of Health (http://www.fnih.org). The grantee organization is the Northern California Institute for Research and Education, and the study is coordinated by the Alzheimer’s Therapeutic Research Institute at the University of Southern California. ADNI data are disseminated by the Laboratory for Neuro Imaging at the University of Southern California. AVID Radiopharmaceuticals, Inc., supplied the AV-1451 precursor, chemistry production advice and oversight; the FDA provided regulatory cross-filing permission and documentation needed for work in the MCSA cohort.

## Author Contributions

KN, SLR, and AJS were involved with study design, statistical analysis, data generation, and drafting of the final manuscript.

PJB was involved with drafting of the final manuscript.

LGA, JB, MRF, TF, KN, PA, RS, BH, SS, DS, CRJ, WJJ, SL, AV, JFW, VD, SML, TP, CCR, VLV, LD, TJH, JL, EM, RFB, KJ, HSY, RCP, VKR, PV, ADC, KHF, MIK, OLL, DAB, MA, TB, CC, DH, PLDJ, MF, VJ, BTL, APT, IC, JM, and MWW were all involved with generation of the imaging, genetic, other “omics,” and/or animal model data.

RD, KF, TYJ, JPK, KN, BH, AV, VD, VLV, SML, TP, LD, TJH, JL, EM, RFB, HSY, ADC, KHF, DAB, VJ, APT, and IC were involved with processing and analysis of imaging and “omics” data.

All authors reviewed and approved the final submitted manuscript.

**Code availability.** The analysis was produced with standard code for software programs utilized, which can be made available from the corresponding author on reasonable request. All software used is freely available online.

## Data availability

Summary statistics will be made available for download upon publication.

**URLs.** PLINK software, https://www.cog-genomics.org/plink/; Michigan Imputation Server, https://imputationserver.sph.umich.edu/index.html#!pages/home; LocusZoom software, http://locuszoom.org/; GSA-SNP software, https://sourceforge.net/projects/gsasnp2/; KGG software, http://pmglab.top/kgg/; SPM12 software, https://www.fil.ion.ucl.ac.uk/spm/software/spm12/; Synapse database, https://www.synapse.org/; eQTLGen Consortium, https://www.eqtlgen.org/; ADNI LONI, https://adni.loni.usc.edu/; ROS/MAP cohort, https://www.radc.rush.edu/; Allen Human Brain Atlas, https://portal.brain-map.org/

