## Supplemental text and figures for "Novel *CYP1B1-RMDN2* Alzheimer’s disease locus identified by genome-wide association analysis of cerebral tau deposition on PET"

### Supplemental Information

#### Table of Contents

Supplemental Methods (Includes Supplemental Tables 1 – 12) – Pages 2 – 24

Supplemental Figure 1 – Page 25

Supplemental Figure 2 – Page 26

Supplemental Figure 3 – Pages 27

Supplemental Figure 4 – Pages 28-29

Supplemental Figure 5 – Pages 30-31

Supplemental Figure 6 – Pages 32-33

Supplemental Figure 7 – Pages 34-35

Supplemental Figure 8 – Pages 36

Supplemental References – Pages 37-45

#### Supplemental Methods

*Discovery sample cohorts*

**Alzheimer's Disease Neuroimaging Initiative (ADNI)**: The ADNI initial phase (ADNI-1) was launched in 2003 to test whether serial magnetic resonance imaging (MRI), position emission tomography (PET), other biological markers, and clinical and neuropsychological assessment could be combined to measure the progression of mild cognitive impairment (MCI) and early AD. ADNI-1 has been extended in subsequent phases (ADNI-GO, ADNI-2, and ADNI-3) for follow-up of existing participants and additional new enrollments. More information about ADNI can be found in previous publications[1-4] and at https://adni.loni.usc.edu. Demographic information, pre-processed tau PET scans, *APOE* and whole-genome genotyping data, neuropsychological test scores, and clinical information are publicly available from the LONI ADNI data repository. A total of 567 ADNI participants (351 cognitively normal older adults (CN), 168 MCI, and 48 AD) were available for analysis. [$^{18}$F]flortaucipir scans were collected using previously described methods (http://adni.loni.usc.edu; [5]). Pre-processed [$^{18}$F]flortaucipir, [$^{18}$F]florbetapir, and [$^{18}$F]florbetaben scans were also downloaded from the ADNI data repository and processed using standard techniques using Statistical Parametric Mapping (SPM12), including normalization to MNI (Montreal Neurological Institute) space. Then, for [$^{18}$F]flortaucipir, standardized uptake value ratio (SUVR) images were created by intensity-normalization using a cerebellar crus reference region. Mean [$^{18}$F]flortaucipir SUVR was extracted from a global cortical grey matter region generated from subject-specific parcellations of structural MPRAGE images generated using FreeSurfer version 6. For [$^{18}$F]florbetaben and [$^{18}$F]florbetapir, scans were processed using the Centiloid (CL) method as previously described[6-8], using local formulas derived from the stage 2 process as described in Risacher et al. 2021[8].

Global cortical CL≥20.76 was considered as Aβ positive (Aβ+), as this cut-off best predicted the SUVR cut-offs produced by University of California - Berkeley (SUVR>1.11 for [$^{18}$F]florbetapir and SUVR>1.08 for [$^{18}$F]florbetaben, *data not shown*). Demographic and neuropsychological performance data for ADNI can be found in **Supplemental Table 1**.

**Supplemental Table 1. ADNI Demographics (Mean (Standard Deviation))**

| | CN (n=351) | MCI (n=168) | AD (n=48) | p-value |
|---|---|---|---|---|
| Age (years) | 73.2 (7.0) | 74.9 (8.1) | 77.4 (9.8) | <0.001 |
| Sex (M, F) | 165, 186 | 97, 71 | 28, 20 | 0.042 |
| Education (years) | 16.7 (2.4) | 16.3 (2.7) | 16.2 (2.7) | 0.088 |
| *APOE* ε4 carrier (%) | 36.5% | 36.3% | 56.3% | 0.026 |
| Aβ Positivity (%)[a] | 54.3% | 56.9% | 66.7% | n.s. |
| FAQ Total Score[b] | 1.4 (4.5) | 3.3 (5.6) | 8.9 (9.6) | <0.001 |
| CDR Global Score[c] | 0.1 (0.3) | 0.4 (0.3) | 0.6 (0.6) | <0.001 |
| CDR Sum of Boxes[c] | 0.5 (1.6) | 1.4 (1.9) | 3.5 (3.6) | <0.001 |
| MMSE Total Score[d,h] | 28.5 (2.4) | 27.6 (3.1) | 25.3 (4.3) | <0.001 |
| ADAS Total Score[e,h] | 6.0 (4.2) | 9.1 (5.4) | 14.8 (9.1) | <0.001 |
| Story Recall Z-Score – Learning[f,i] | 0.0 (1.1) | -0.6 (1.2) | -1.1 (1.5) | <0.001 |
| Story Recall Z-Score – Delayed[f,i] | 0.0 (1.2) | -0.7 (1.2) | -1.3 (1.6) | <0.001 |
| Trail Making – Part A[g,i] | 32.4 (12.4) | 36.8 (18.9) | 43.8 (26.2) | <0.001 |
| Trail Making – Part B[g,i] | 84.5 (55.7) | 98.9 (62.4) | 133.1 (61.8) | 0.001 |
| Animal Fluency[h,i] | 20.8 (6.2) | 18.7 (5.8) | 17.1 (6.6) | 0.002 |

[a] 2 participants (1 CN, 1 MCI) missing Aβ positivity
[b] 21 participants (11 CN, 8 MCI, 2 AD) missing FAQ score
[c] 19 participants (10 CN, 6 MCI, 3 AD) missing CDR scores
[d] 13 participants (8 CN, 3 MCI, 2 AD) missing MMSE score
[e] 85 participants (47 CN, 29 MCI, 9 AD) missing ADAS score
[f] 13 participants (6 CN, 5 MCI, 2 AD) missing Story Recall z-scores (*z-score calculated relative to CN individuals)
[g] 20 participants (8 CN, 9 MCI, 3 AD) missing Trail Making Test scores
[h] 12 participants (5 CN, 5 MCI, 2 AD) missing Animal Fluency
[i] Covaried for age, sex, and years of education

**ADNI-Department of Defense (ADNI-DoD)**: ADNI-DoD is an extension of the original ADNI (described above) to investigate the relationship of traumatic brain injury (TBI) and post-traumatic stress disorder (PTSD) on the development of AD. ADNI-DoD aims to enroll approximately 400 elderly Veterans who served in Vietnam, including some individuals with PTSD and/or TBI, as well as controls. Individuals in the study are diagnosed as CN or MCI using ADNI criteria (http://www.loni.usc.edu/ADNI). Participants in ADNI-DoD undergo all procedures that are collected as part of the original ADNI, including cognitive tests and clinical examinations, amyloid PET with [$^{18}$F]florbetapir, structural and functional MRI, lumbar puncture for CSF analysis, and blood collection for genetic analysis, including *APOE* and genome-wide data. In addition, a subset of participants also underwent tau PET imaging with [$^{18}$F]flortaucipir. A total of 68 participants from ADNI-DoD (55 CN, 13 MCI) were included in the present analysis. For more information, see a previous report from Weiner et al. (2017)[9]. [$^{18}$F]flortaucipir scans from ADNI-DoD were processed using an identical processing pipeline to that in the ADNI study. Specifically, [$^{18}$F]flortaucipir scans were collected using previously described methods (http://adni.loni.usc.edu)[5]. Pre-processed [$^{18}$F]flortaucipir scans were downloaded from the ADNI data repository (http://adni.loni.usc.edu) and processed using standard techniques using Statistical Parametric Mapping (SPM12), including normalization to MNI (Montreal Neurological Institute) space. Then, standard uptake value ratio (SUVR) images were created by intensity-normalization using a cerebellar crus reference region. Mean [$^{18}$F]flortaucipir SUVR was extracted from a global cortical grey matter region generated from subject-specific parcellations of structural MPRAGE images generated using from FreeSurfer version 6. As in ADNI, amyloid scans were also collected for 67 participants (1 MCI missing amyloid data) using only [$^{18}$F]florbetapir. Briefly, static images were downloaded from the ADNI data repository (http://adni.loni.usc.edu) and processed using standard methods, including normalization to MNI space, in SPM12. Again, scans were processed using the CL method as described above and previously[6-8], and CL $\geq$ 20.76 was consider A$\beta$+. Demographic and neuropsychological performance data for ADNI-DoD can be found in **Supplemental Table 2**.

| **Supplemental Table 2. ADNI-DoD Demographics (Mean (Standard Deviation))** | | | |
|---|---|---|---|
| | CN (n=55) | MCI (n=13) | p-value |
| Age (years) | 71.1 (5.6) | 70.1 (2.5) | n.s. |
| Sex (M, F) | 54, 1 | 13, 0 | n.s. |

| | | | |
|---|---|---|---|
| Education (years) | 15.4 (2.5) | 15.0 (2.9) | n.s. |
| *APOE* ε4 carrier (%) | 25.5% | 15.4% | n.s. |
| Aβ Positivity (%)[a] | 23.6% | 25.0% | n.s. |
| FAQ Total Score[b] | 1.3 (2.5) | 4.8 (5.7) | 0.012 |
| CDR Global Score[c] | 0.1 (0.2) | 0.4 (0.2) | <0.001 |
| CDR Sum of Boxes[c] | 0.3 (0.7) | 1.3 (1.3) | <0.001 |
| MMSE Total Score[f] | 28.4 (1.8) | 27.5 (2.6) | n.s. |
| ADAS-11 Total Score[f] | 6.4 (3.5) | 8.5 (3.6) | *0.090* |
| Story Recall Z-Score – Learning[d,f] | 0.0 (1.1) | -1.1 (1.2) | 0.003 |
| Story Recall Z-Score – Delayed[d,f] | 0.1 (1.1) | -1.2 (1.0) | <0.001 |
| Trail Making – Part A[e,f] | 36.7 (17.8) | 49.4 (33.6) | <0.001 |
| Trail Making – Part B[e,f] | 82.4 (27.4) | 150.8 (84.6) | <0.001 |
| Animal Fluency[f] | 20.9 (5.0) | 17.2 (5.6) | 0.023 |
| [a] 1 MCI participant missing Aβ positivity<br>[b] 31 participants (28 CN, 3 MCI) missing FAQ score<br>[c] 9 CN participants missing CDR scores | [d] 1 CN participant missing Trail Making Test scores<br>[e] z-score calculated relative to CN individuals<br>[f] Covaried for age, sex, and years of education | | |

**<u>Indiana Memory and Aging Study (IMAS)</u>**: IMAS is a longitudinal observational study of older adults at risk for and with prodromal and clinical AD. As part of the IMAS study, participants underwent a comprehensive clinical visit through the Indiana Alzheimer's Disease Research Center (IADRC), which included clinical and neurologic testing, neuropsychological testing with the Uniform Dataset 3 (UDS-3) battery, and blood samples, as well as structural and functional MRI, [$^{18}$F]flortaucipir PET scans, and amyloid PET scans with either [$^{18}$F]florbetapir or [$^{18}$F]florbetaben. AD and MCI were diagnosed using standard criteria[10,11]. CN were older adults without a significant performance deficit on cognitive testing. A total of 54 participants from IMAS (30 CN, 15 MCI, 9 AD) were included in the present analysis. In IMAS, [$^{18}$F]flortaucipir scans were collected as previously described[7,8]. Using SPM12, the middle four 5-minute frames (80-100 minutes) were motion corrected, normalized to MNI space, averaged to create an 80-100 minute static image, intensity normalized to the cerebellar crus to create SUVR images, and smoothed with an 8mm FWHM (Full Width at Half Maximum) Gaussian kernel. Mean [$^{18}$F]flortaucipir SUVR was extracted from a global cortical grey matter region generated from subject-specific parcellations of structural MPRAGE images generated using from FreeSurfer version 6. Amyloid scans were also collected for 55 individuals in IMAS (3 MCI missing amyloid data) using both [$^{18}$F]florbetapir and [$^{18}$F]florbetaben in IMAS as previously described[7,8]. Using SPM12, the four 5-minute frames were spatially aligned to each participant's individual magnetization-prepared rapid gradient-echo (MPRAGE) scan, motion corrected, normalized to Montreal Neurologic Institute (MNI) space, and averaged to create a 50-70 minute static image for the [$^{18}$F]florbetapir or a 90-110 minute static image for the [$^{18}$F]florbetaben. Then SUVR images were created by intensity-normalizing to the whole cerebellum for both tracers. The whole cerebellum ROI was from the CL project. The resulting SUVR images were converted to CL units and smoothed using an 8mm full-width half maximum (FWHM) Gaussian kernel. Again, CL ≥ 20.76 was consider Aβ+. Demographic and neuropsychological performance data for IMAS can be found in **Supplemental Table 3**.

**Supplemental Table 3. IMAS Demographics (Mean (Standard Deviation))**

| | CN (n=30) | MCI (n=15) | AD (n=9) | p-value |
|---|---|---|---|---|
| Age (years) | 70.1 (7.7) | 72.6 (8.3) | 64.4 (11.2) | *0.083* |
| Sex (M, F) | 8, 22 | 8, 7 | 4, 5 | n.s. |
| Education (years) | 17.0 (2.2) | 15.6 (3.3) | 15.6 (2.6) | n.s. |
| *APOE* ε4 carrier (%) | 40.0% | 66.7% | 55.6% | n.s. |
| Aβ Positivity (%)[a] | 23.3% | 41.7% | 77.8% | 0.012 |
| FAQ Total Score | 0.2 (0.6) | 4.5 (4.4) | 10.8 (6.5) | <0.001 |
| CDR Global Score | 0.1 (0.2) | 0.5 (0.1) | 0.7 (0.5) | <0.001 |
| CDR Sum of Boxes | 0.1 (0.3) | 1.9 (1.2) | 4.7 (3.0) | <0.001 |
| MMSE Total Score[b,e,*] | 29.6 (0.8) | 26.3 (3.2) | 21.6 (5.1) | <0.001 |
| Story Recall Z-Score – Learning[c,i] | 0.0 (1.0) | -1.4 (0.9) | -2.3 (0.5) | <0.001 |
| Story Recall Z-Score – Delayed[c,i] | 0.0 (1.0) | -2.3 (0.9) | -2.9 (0.6) | <0.001 |
| Trail Making – Part A[d,e] | 26.2 (9.2) | 34.9 (11.3) | 44.9 (14.5) | <0.001 |
| Trail Making – Part B[d,e] | 65.5 (24.7) | 107.1 (35.7) | 182.9 (64.2) | <0.001 |
| Animal Fluency[e] | 25.0 (4.3) | 16.8 (4.6) | 11.3 (5.5) | <0.001 |

[a] 3 MCI participants missing Aβ positivity
[b] 1 MCI participant missing MMSE score
[c] 1 AD participant missing Story Recall z-scores (*z-score calculated relative to CNs)
[d] 2 AD participants missing Trail Making Test scores
[e] Covaried for age, sex, and years of education
* imputed from MoCA scores using [12]

**<u>A05 Study</u>**: 169 participants (51 CN, 74 MCI, 44 AD) were recruited at 25 sites and underwent cognitive testing and clinical assessment, structural MRI, amyloid PET with [$^{18}$F]florbetapir, tau PET with [$^{18}$F]flortaucipir, and blood collection for genetic analysis, including *APOE* and genome-wide data[13,14]. Structural MRIs, tau scans, and genome-wide data were provided by Avid/Eli Lilly under a research agreement, while clinical and cognitive data, *APOE* genotype (2 CN, 3 MCI missing *APOE* genotype data), and Aβ positivity (missing for 1 AD patient) were provided in a summary dataset. Tau PET scans were provided as 80-100 minute static images, which were subsequently co-registered to structural MRI scans from the same visit and normalized to MNI space using parameters from segmentation of the structural MRIs using SPM12. Then, standard uptake value ratio (SUVR) images were created by intensity-normalization using a cerebellar crus reference region. Mean [$^{18}$F]flortaucipir SUVR was extracted from a global cortical grey matter region generated from subject-specific parcellations of structural MPRAGE images generated using from FreeSurfer version 6. Demographic and neuropsychological performance data for A05 can be found in **Supplemental Table 4**.

| **Supplemental Table 4. A05 Demographics (Mean (Standard Deviation))** | | | | |
|---|---|---|---|---|
| | CN (n=51) | MCI (n=74) | AD (n=44) | p-value |
| Age (years) | 63.6 (18.2) | 71.6 (9.1) | 74.0 (9.0) | <0.001 |
| Sex (M, F) | 29, 22 | 41, 33 | 20, 24 | n.s. |
| *APOE* ε4 carrier (%) | 19.6% | 48.6% | 54.5% | <0.001 |
| Aβ Positivity (%)[a] | 9.8% | 50.0% | 65.1% | <0.001 |
| FAQ Total Score[b] | n/a | 4.3 (4.9) | 15.2 (6.9) | <0.001 |
| MMSE Total Score[e] | 29.6 (0.5) | 27.9 (1.8) | 22.0 (4.0) | <0.001 |
| ADAS Total Score[e] | 5.4 (3.1) | 10.1 (4.4) | 19.9 (8.7) | <0.001 |
| Story Recall Z-Score – Learning[c,e] | 0.0 (1.0) | -1.0 (0.8) | -2.0 (-0.8) | <0.001 |
| Story Recall Z-Score – Delayed[c,e] | 0.0 (1.0) | -1.0 (0.8) | -2.1 (0.5) | <0.001 |
| Trail Making – Part A[d,e] | 35.2 (15.1) | 48.2 (21.4) | 73.2 (43.6) | <0.001 |
| Trail Making – Part B[d,e] | 86.7 (42.8) | 126.5 (77.3) | 204.4 (101.2) | <0.001 |
| Animal Fluency[e] | 21.6 (6.3) | 16.8 (4.7) | 9.6 (4.6) | <0.001 |

| | |
|---|---|
| [a] 1 AD participants missing Aβ positivity<br>[b] All CN participants (n=51) missing FAQ score<br>[c] Z-score calculated relative to CN individuals | [d] 3 AD participants missing Trail Making Test scores<br>[e] Covaried for age, sex, and years of education |

**A4 Study**: The A4 trial is an ongoing 240-week pharmacological trial testing whether an anti-amyloid treatment, solanezumab, can slow cognitive decline and progression to dementia in older adults with preclinical AD (i.e., amyloid-positive cognitively normal older adults)[15,16]. Participants were recruited and assessed at 67 clinical trial sites across the world and included in the study if they were positive on an amyloid PET scan with [$^{18}$F]florbetapir, cognitively normal, and met all other study criteria. A subset of 303 CN participants, including 248 participants from the main A4 study and 55 participants from part of the LEARN sub-study, received a tau PET scan with [$^{18}$F]flortaucipir that was downloaded as 80-100 minute status on the LONI data repository (https://ida.loni.usc.edu/home/projectPage.jsp?project=A4). Then, the [$^{18}$F]flortaucipir scans were co-registered to structural MRI scans from the same visit and normalized to MNI space using parameters from segmentation of the structural MRIs using SPM12. Standard uptake value ratio (SUVR) images were created by intensity-normalization using a cerebellar crus reference region, and mean [$^{18}$F]flortaucipir SUVR was extracted from a global cortical grey matter region generated from subject-specific parcellations of structural MPRAGE images generated using from FreeSurfer version 6. Amyloid scans were also collected for all participants using only [$^{18}$F]florbetapir. Briefly, static images were downloaded from the A4 data repository (http://adni.loni.usc.edu) and processed using standard methods, including normalization to MNI space, in SPM12. Again, scans were processed using the CL method as described above and previously[6-8] and CL ≥ 20.76 was consider Aβ+. Demographic and neuropsychological performance data for A4 can be found in **Supplemental Table 5**.

| **Supplemental Table 5. A4 Demographics (Mean (Standard Deviation))** | |
|---|---|
| | CN (n=303) |
| Age (years) | 71.6 (4.7) |
| Sex (M, F) | 185, 118 |
| Education (years) | 16.3 (2.7) |
| *APOE* ε4 carrier (%) | 54.5% |
| Aβ Positivity (%) | 86.8% |
| CDR Global Score | 0.0 (0.0) |
| CDR Sum of Boxes | 0.1 (0.2) |
| MMSE Total Score | 28.7 (1.3) |
| Story Recall Z-Score – Learning | -0.1 (0.8) |
| Story Recall Z-Score – Delayed | -0.1 (0.8) |

*Genotyping and Imputation for ADNI, ADNI-DOD, A05, A4, and IMAS*

Participants were genotyped using several Illumina genotyping platforms. As the cohorts used different genotyping platforms, un-genotyped SNPs were imputed separately in each cohort using MACH and the Haplotype Reference Consortium (HRC) data as a reference panel[17]. Before the imputation, standard sample and SNP quality control procedures were performed as described previously[18]: (1) for SNP, SNP call rate < 95%, Hardy-Weinberg $p$-value<$1\times10^{-6}$, and minor allele frequency (MAF) < 1%; (2) for sample, sex inconsistencies, and sample call rate < 95%. Furthermore, in order to prevent spurious association due to population stratification, only non-Hispanic participants of European ancestry that clustered with HapMap CEU (Utah residents with Northern and Western European ancestry from the CEPH collection) or TSI (Toscani in Italia) populations were selected using multidimensional scaling (MDS) analysis (www.hapmap.org)[19]. Imputation and quality control procedures were performed as described previously[20]. After the imputation, an $r^2$ value equal to 0.30 was used as the threshold to accept the imputed genotypes.

**Harvard Aging Brain Study (HABS):** HABS is a longitudinal study of aging and dementia that includes more than 300 older adults who were cognitively normal at enrollment. More information about HABS can be found in previous publications[21-24]. A subset of 147 individuals (139 CN, 6 MCI, 2 dementia) from HABS had GWAS, cognitive, *APOE* genotype, and [$^{18}$F]flortaucipir PET data and were included in this analysis. HABS [$^{18}$F]flortaucipir PET scan acquisition parameters and processing protocols have been published previously (Johnson et al 2016, Dagley et al. 2017, Buckley et al. 2018). SUVR images were generated from 80 to 100 minutes static images. Global cortical grey matter [$^{18}$F]flortaucipir SUVR was extracted and used at the phenotype for GWAS analysis. Demographic and neuropsychological performance data for HABS can be found in **Supplemental Table 6**.

In HABS, genome-wide genotyping was performed in two batches, using the same genotyping platform (Axiom™ Biobank Genotyping Array): the first batch (n=189) was genotyped in 2013, while the second batch (n=96) was completed in 2016. We selected single nucleotide polymorphisms (SNPs) and individuals according to the following quality control criteria, using PLINK v1.9: autosomal SNPs with minor allele frequency (MAF)>0.01, genotype missing rate (variant)<0.02, genotype missing rate (sample)<0.02, non-extreme heterozygosity (within 5 s.d.), identity by descent (IBD) pi-hat<0.125, and Hardy-Weinberg equilibrium (HWE) $p>10^{-6}$. We selected participants of European descent using multi-dimensional scaling (MDS), resulting in 155 participants with 290,073 variants from the 2013 batch, and 61 participants with 264,623 variants from the 2016 batch. These two data files were merged only taking matching SNPs across two batches, resulting in 216 participants with 239,503 variants. PLINK v1.9 was used to derive genotype principal components that capture population structures. Then, Eagle v2.4 was used to phase the genotypes, and imputation was done through Minimac4 on Michigan

Imputation Server using the Haplotype Reference Consortium (HRC) reference panel (r1.1 2016, GRCh37/hg19). After post-imputation QC (MAF>0.01, HWE $p>10^{-6}$, and imputation $R^2>0.9$), we had 5,673,786 autosomal variants from 216 individuals of European ancestry (of which 150 were used in this analysis). Finally, a regression model implemented in PLINK was calculated and summary association values for all variants were shared with IUSM.

| **Supplemental Table 6. HABS Demographics (Mean (Standard Deviation))** | | |
|---|---|---|
| | HABS CN (n=139) | HABS MCI (n=6) |
| Mean age, years (sd) | 79.3 (6.2) | 80.3 (6.8) |
| Female (%) | 79 (57) | 3 (50) |
| Mean Education, years (sd) | 16.6 (3.0) | 16 (1.3) |
| *APOE* ε4 carrier (%)** | 35 (25) | 5 (83) |
| MMSE, median (IQR) | 29 (1) | 27 (2.3) |

*Two dementia cases and three cases with missing diagnoses whose demographics are not summarized to protect participant privacy
**One CN participant with missing *APOE* genotype info

**University of Pittsburgh ADRC:** Participants were recruited through neuroimaging studies affiliated with the University of Pittsburgh ADRC from studies on cardiovascular risk factors and population-based cohorts from small towns near Pittsburgh and surrounding areas [25]. 138 individuals (59 CN, 35 MCI/AD, 44 Other) with GWAS data and [$^{18}$F]flortaucipir scans were included in the present study. Briefly, [$^{18}$F]flortaucipir scans were processed using standard techniques and global cortical tau SUVR was extracted[25]. Genotype data generated from the Illumina Infinium Global Diversity Array (GDA) underwent standard QC and imputed using HRC. Finally, a regression model implemented in PLINK was calculated and summary association values were shared with IUSM. Demographic and neuropsychological performance data for the University of Pittsburgh ADRC can be found in **Supplemental Table 7**.

| **Supplemental Table 7. UPitt ADRC Demographics (Mean (Standard Deviation))** | | | | |
|---|---|---|---|---|
| | CN (n=59) | MCI/AD (n=35) | Other (n=44) | p-value |
| Age (years) | 72.5 (4.5) | 74.5 (3.8) | 74.4 (5.5) | 0.054 |

| | | | | |
|---|---|---|---|---|
| Sex (M, F) | 18, 41 | 12, 23 | 14, 30 | n.s. |
| Education (years) | 16.3 (2.6) | 14.9 (2.8) | 15.3 (2.8) | 0.048 |
| *APOE* ε4 carrier (%) | 33.9% | 22.9% | 18.2% | n.s. |
| MMSE Total Score[a] | 28.5 (1.4) | 27.3 (2.2) | 27.4 (2.2) | 0.041 |
| CDR Global Score | 0.01 (0.07) | 0.23 (0.25) | 0.10 (0.23) | <0.001 |
| [a]6 participants (2 CN, 4 Other) missing MMSE score | | | | |

*Replication Cohorts*

**Mayo Clinic Study of Aging (MCSA):** The MCSA is a community-based study of older adults (aged 70-89) from Rochester, Minnesota and surrounding regions.[26,27] 649 individuals (571 CN, 66 MCI, 17 AD) with genetic data and [$^{18}$F]flortaucipir scans were included in this analysis. Briefly, [18F]flortaucipir scans were collected according to previously described protocols and processed using standard techniques[28-30]. Regional uptake was measured with an in-house automated pipeline with ROI labels propagated from a custom MRI template[31,32]. Global tau burden was calculated from a composite meta-ROI as previously described[28-30]. GWAS data was generated for all participants using the Illumina Infinium Global Screening Array-24 v2.0 and underwent standard QC using PLINK version 1.9 as previously described[28,33]. GWAS data then underwent imputation using the HRC reference panel, and after QC, 6,417,232 SNPs were available for analysis[28]. The replication analysis was calculated using a linear regression model, covaried for age, sex, ancestry principal components (PCs), APOE genotype, and diagnosis for the top SNPs identified from the discovery cohorts. Demographic and neuropsychological performance data for MCSA participants can be found in **Supplemental Table 8**.

| **Supplemental Table 8. MCSA Demographics (Mean (Standard Deviation))** | | | |
|---|---|---|---|
| | CN (n=571) | MCI (n=66) | AD (n=17) |
| Age (years) | 71.1 (10.5) | 77.1 (9.3) | 83.0 (4.8) |
| Sex (M, F) | 313, 258 | 38, 28 | 10, 7 |

| | | | |
|---|---|---|---|
| Education (years) | 15.0 (2.3) | 13.4 (3.1) | 13.5 (3.0) |
| *APOE* ε4 carrier (%) | 28.1% | 34.8% | 58.8% |

**Memory and Aging Project (MAP) at the Knight ADRC:** For this study, data was collected from different AD-related cohorts enrolled in the MAP at the Knight ADRC. The total sample size included was 309 (279 CN, 30 MCI/AD). Collection of genotype data and PET image processing are described in detail in the respective studies[34-37]. Briefly, participants were diagnosed as cognitively normal (controls) or AD (cases), based on CDR global score. For each cohort, tau PET SUVR was extracted from a composite of cortical brain areas[34,38]. The normalized z-score SUVRs were calculated by using the mean and standard deviation (SD) units for all the samples and applied to the entire cohort. Stringent QC steps of the GWAS data were applied. Briefly, we used the threshold of 98% for removing single nucleotide polymorphisms (SNPs) and participants with low call rate. Autosomal SNPs that were not in the Hardy-Weinberg equilibrium ($P<1\times10-6$) were also removed. Duplication and relatedness of participants were estimated from identity-by-descent (IBD) analysis carried out in Plink version 1.9[39]. In case of related participants (Pihat ≥ 0.25), the samples with a higher number of variants that passed the QC were prioritized. For phasing and imputation, we used the 1000 Genomes Project Phase 3 data (October 2014), SHAPEIT v2.r837[40], and IMPUTE2 v2.3.2[41,42]. We used imputed probability score<0.90 and ≥ 0.90 as thresholds for missing and fully observed participant genotypes, respectively. Genotyped and imputed variants with MAF<0.02 or IMPUTE2 information score<0.30 were discarded. Principle component analysis (PCA) was performed on the genotype data to obtain genetic PCs that capture population substructure. Statistical analyses and data visualization were performed in Plink version 1.9[43] and R version 3.5.2. We performed association analyses of the strongest variants found in the discovery cohort with tau pathology. The associations were tested using logistic regression model from the base R “stats” package, adjusted for sex, age, *APOE* genotype, diagnosis, and the first two genetic PCs. Demographic and neuropsychological performance data for MAP participants can be found in **Supplemental Table 9**.

| **Supplemental Table 9. MAP at the Knight ADRC Demographics (Mean (Standard Deviation))** | | |
|---|---|---|
| | CN (n=279) | MCI/AD (n=30) |
| Age (years) | 69.8 (8.1) | 77.4 (7.0) |
| Sex (M, F) | 128, 184 | 16, 16 |
| Education (years) | 16.3 (2.3) | 15.4 (3.0) |
| *APOE* ε4 carrier (%) | 33.3% | 62.5% |

**Australian Imaging, Biomarker and Lifestyle Study (AIBL):** The AIBL study is a longitudinal cohort study collected at multiple sites in Australia[44,45]. A subset of AIBL individuals had genetic data and tau PET imaging and thus, were included in this analysis, including 77 individuals (65 CN, 8 MCI, 4 AD) who underwent tau PET with [$^{18}$F]flortaucipir and 458 who underwent tau PET with [$^{18}$F]MK-6240. Briefly, [18F]flortaucipir and [18F]MK-6240 scans were collected as a 20-minute acquisition either 80 minutes or 90 minutes post-injection of tracer, respectively. Scan processing was done using standard methods to generate SUVR images[46]. Finally, mean global cortical [18F]flortaucipir and [18F]MK-6240 SUVR was extracted. GWAS SNP array data with 976,713 SNPs (including 273,000 exome variants and an additional 13,000 custom content SNPs) on the OmniExpressHumanExome+BeadChip (Illumina, USA) was generated. Genetic data was mapped to the human genome reference hg19 and standard QC was performed as previously described (Fowler et al. 2021). Finally, the dataset was imputed to the 1000 Genomes Project Phase 3. Linear regression models testing the top SNPs identified in the discovery cohort were calculated separately for the [$^{18}$F]flortaucipir and [$^{18}$F]MK-6240 cohorts, covaried for age, sex, *APOE* genotype, diagnosis, and ancestry PCs. Demographic and neuropsychological performance data for AIBL participants who underwent

[$^{18}$F]flortaucipir scans can be found in **Supplemental Table 10**, while demographic and neuropsychological performance data for participants who underwent [$^{18}$F]MK-6240 scans can be found in **Supplemental Table 11**.

**Supplemental Table 10. AIBL [$^{18}$F]Flortaucipir Cohort Demographics (Mean (Standard Deviation))**

| | CN (n=66) | MCI (n=8) | AD (n=4) |
|---|---|---|---|
| Age (years) | 75.0 (7.0) | 74.0 (6.0) | 75.0 (8.0) |
| Sex (M, F) | 32, 34 | 5, 3 | 2, 2 |
| Education (years) | 12.5 (3.0) | 9.5 (6.0) | 14.5 (5.0) |
| Aβ Positivity (%) | 39.0% | 50.0% | 75.0% |
| MMSE Total Score | 28.0 (1.0) | 26.5 (2.0) | 22.5 (5.0) |
| CDR Global Score | 0.0 (0.1) | 0.5 (0.2) | 0.8 (0.5) |
| CDR Sum of Boxes | 0.1 (0.3) | 1.1 (1.1) | 4.2 (3.0) |

**Supplemental Table 11. AIBL [$^{18}$F]MK-6240 Cohort Demographics (Mean (Standard Deviation))**

| | CN (n=271) | MCI (n=102) | AD (n=85) |
|---|---|---|---|
| Age (years) | 76.0 (6.0) | 74.0 (8.0) | 71.0 (8.0) |
| Sex (M, F) | 123, 156 | 61, 44 | 45, 40 |
| Education (years) | 14.0 (3.0) | 12.0 (3.0) | 12.0 (3.0) |
| Aβ Positivity (%) | 19.0% | 58.0% | 83.0% |
| MMSE Total Score | 28.5 (1.0) | 26.5 (2.0) | 22.5 (4.0) |
| CDR Global Score | 0.0 (0.1) | 0.5 (0.1) | 0.8 (0.5) |
| CDR Sum of Boxes | 0.1 (0.3) | 1.3 (0.9) | 5.2 (2.5) |

**Berkeley Aging Cohort Study (BACS):** The BACS began recruiting cognitively normal individuals from the community in 2005[47,48]. 107 individuals with genotyping and [$^{18}$F]flortaucipir were included in the analysis. [$^{18}$F]flortaucipir scans were collected as previously described and processed using standard techniques to generate SUVR images. Global cortical [$^{18}$F]flortaucipir SUVR was extracted for each scan using Freesurfer-generated ROIs. Genotype data was generated using the Illumina OmniExpress Exome panel and subjected to standard QC[49]. Data was then imputed using European samples from the HRC r1.1.216 reference panel (Build 37, Assembly 19) and filtered for quality and minor allele frequency. Finally, linear regression models were calculated to assess the relationship of the top SNPs from the discovery cohorts to tau deposition, covaried for age, sex, *APOE* genotype, and the first two ancestry PCs. Demographic and neuropsychological performance data for BACS participants who underwent [$^{18}$F]flortaucipir scans can be found in **Supplemental Table 12.**

| **Supplemental Table 12. BACS Demographics (Mean (Standard Deviation))** | |
|---|---|
| | CN (n=107) |
| Age (years) | 70.8 (11.8) |
| Sex (M, F) | 45, 63 |
| Education (years) | 16.8 (1.8) |
| *APOE* ε4 carrier (%) | 31.0% |
| MMSE Total Score | 29.0 (1.1) |

**Gene-set enrichment analysis:** Gene-set enrichment analysis was performed using GWAS summary statistics to identify pathways and functional gene sets with significant associations with global cortical tau deposition levels using the GSA-SNP software, as described in the Supplemental information. The GSA-SNP uses a p-value of each SNP from GWAS summary statistics to test if a pathway-phenotype association is significantly different from all other pathway-phenotype associations[50,51]. Gene-set enrichment analysis was restricted to pathways containing between 10 and 200 genes. False discovery rate (FDR) with the Benjamini-Hochberg procedure was used for multiple comparison correction[52]. Significantly enriched pathways with global cortical tau levels were defined as those with FDR-corrected $p$-value<0.05.

**Gene-based association analysis:** Genome-wide gene-based association analysis was performed using GWAS p-values and the KGG software as described previously[51,53]. First, SNPs in each gene were divided into different LD blocks depending on pairwise LD coefficients ($r^2$) for all SNPs. Second, for each block, a block-based p-value for association was calculated, and the key SNP was derived and marked. Next, the block-based p-values were combined, accounting for LD between the key SNPs using the scaled chi-square.

**<u>Interaction with diagnosis, *APOE* genotype, and amyloid positivity</u>:** Using data from ADNI, ADNI-DOD, IMAS, A4, and A05, we assessed the effect of the identified SNP (rs2113389 – dominant model (CC vs CT/TT)) and its interaction with other variables of interest on global and regional tau (i.e., medial temporal lobe (MTL) tau, the primary location of early tau deposition in the early stages of AD). 1154 individuals, including 789 CN, 265 MCI, and 100 AD, were included in these *post-hoc* analyses (12 participants (3 CN, 8 MCI, 1 AD) were excluded for missing either *APOE* genotype or amyloid status data). The effect of the identified SNP on MTL and global tau (both rank-based inverse normal transformed) was assessed using an ANCOVA model, covaried for age, sex, *APOE* ε4 carrier status, amyloid positivity, diagnosis, and the first two population PCs. Next, we tested whether there was an interaction between the identified SNP (rs2113389 – dominant model (CC vs CT/TT)) with diagnosis on MTL, and global tau was evaluated using a two-way ANCOVA model, covaried for age, sex, *APOE* ε4 carrier status, amyloid positivity, and the first two population PCs. Similarly, the interaction effect of the identified SNP (rs2113389 – dominant model (CC vs CT/TT)) with *APOE* ε4 carrier status on MTL and global tau was evaluated using a two-way ANCOVA model, covaried for age, sex, amyloid positivity, diagnosis, and the first two population PCs. Finally, the interaction effect of the identified SNP (rs2113389 – dominant model (CC vs CT/TT)) with amyloid positivity status on MTL and global tau was evaluated using a two-way ANCOVA model, covaried for age, sex, *APOE* ε4 carrier status, diagnosis, and the first two population PCs.

In addition, we sought to determine whether differential effects were observed in a sex-stratified analysis, given previous studies suggesting that females have higher tau deposition than males. Thus, we tested the main effect of the identified SNP (rs2113389 – dominant model (CC vs

CT/TT)) on MTL and global tau, as well as its interaction with diagnostic group, *APOE* ε4 carrier status, and amyloid positivity status, in males (n=648) and females (n=506) separately.

**Detailed whole-brain imaging analysis:** The tau PET SUVR images were used in a voxel-wise statistical analysis of the effect of the top identified SNP on tau deposition using SPM12 (www.fil.ion.ucl.ac.uk/spm/) in a *post-hoc* analysis (n=1154). A multivariable regression analysis, masked for grey and white matter, was performed using age, sex, *APOE* ε4 carrier status, amyloid positivity, diagnosis, and the first two ancestry PCs as covariates. In the voxel-wise whole brain analysis, a voxel-wise threshold of p<0.05 with family-wise error (FWE) adjustment for multiple comparisons was used. Both an additive and dominant model for the identified SNP (rs2113389 – dominant model (CC vs CT/TT)) were tested.

**AMP-AD bulk RNA-Seq data in the post-mortem human brain:** Pre-processed bulk RNA-Seq data were downloaded from the Synapse database (https:// www. synapse.org/) of the Accelerating Medicines Partnership for Alzheimer's Disease (AMP-AD) Consortium[54-58]: the Mayo Clinic Brain Bank, the Mount Sinai Medical Center Brain Bank (MSBB), and the Religious Orders Study and Memory and Aging Project cohorts (ROSMAP), where RNA-Seq data were generated from the temporal cortex and cerebellum, from parahippocampal gyrus, inferior frontal gyrus, superior temporal gyrus, and frontal pole, and from dorsolateral prefrontal cortex, respectively. The procedures for sample collection, post-mortem sample descriptions, tissue and RNA preparation methods, library preparation and sequencing methods, and sample quality controls were previously described in detail[58].

**Single-nucleus RNA-Seq (snRNA-Seq) analysis:** Single-nucleus RNA-Seq preprocessing: Frozen brain tissue specimens (N=479) from the dorsolateral prefrontal cortex (DLFPC) were obtained in the ROSMAP cohort[59]. Nuclei suspension was prepared as described previously[60]. 5,000 nuclei from each of eight participants were then pooled into one sample, and the 40,000 nuclei in around 15-30 ul volume were loaded into the 10X Single Cell RNA-Seq Platform using the Chromium Single Cell 3' Reagent Kits version 3. Libraries were made following the manufacturer's protocol and sequenced on using Illumina NovaSeq 6000. All raw data are available through the AD Knowledge Portal (https://www.synapse.org/#!Synapse:syn31512863). FASTQ files were processed using the CellRanger software (10x Genomics). The "remove-background" module of CellBender (https://github.com/broadinstitute/CellBender) was used to call cells and eliminate UMI counts of ambient RNA. Our snRNA-Seq library consisted of nuclei from eight individuals. To assign back each nucleus to its participant of origin, each nucleus' genotype data obtained from the snRNA-Seq reads were compared with whole genome sequencing (WGS) SNPs of 1,196 ROS/MAP individuals using demuxlet software[61]. Among 479 specimens analyzed by snRNA-Seq, we excluded the following: specimens that failed in the genotype-based donor assignment, specimens that potentially experienced sample swaps, specimens that had quality problems in WGS, specimens with shallow sequencing depth, and specimens from duplicated individuals. After these quality control processes, 424 individuals remained. Single-nucleus RNA-Seq analysis: Using the cell-type annotations of our previous work[60], we constructed a regularized logistic-regression classifier that infers cell type based on single-nucleus gene expression. This classifier was applied to each cell of single-nucleus libraries. Then, low-quality nuclei were determined using cell-type specific thresholds of total number of UMIs and number of unique features, and low-quality nuclei were removed. Doublets were detected by running DoubletFinder[62]. Pseudobulk UMI count matrix for each cell type was generated by extracting UMI counts of the cell type and by aggregating the counts per gene per individual. Low expression genes were filtered out by using filterByExpr function of edgeR with its default parameters. The pseudobulk counts were normalized by using the trimmed mean of M-values (TMM) method of edgeR, and $\log_2$ of counts per million mapped reads (CPM) were computed using the voom function of limma. Low expression genes whose $\log_2$CPM were less than 2.0 were filtered out. Batch effects were corrected using ComBat.

**Allen Human Brain Atlas data and analysis:** Regional gene expression profiles for 20,736 protein-coding genes were derived from brain-wide microarray-based transcriptome data from the Allen Human Brain Atlas[63,64]. The microarray probes were collected from 3,700 regional brain tissue samples from autopsy data of six adult individuals (5 males/1 female; aged 24–57 years) without history of neurological disorders. Since the first two donors (1 male and 1 female) did not show interhemispheric asymmetries or sex-related differences in gene expression data,[63] the later four donors (all males) had brain tissue collection only in the left hemisphere.[64] The Desikan-Killiany (D-K) atlas was used to parcellate each gene expression map into 68 cortical ROIs.[65] The expression values from multiple probes were mean averaged for each gene. The averaged gene expression values from all voxels in each ROI were mapped into a specific cortical region. Median values were calculated for each gene and each ROI.[66] Here, we focus on the spatial distributions of gene expression profiles in the left hemisphere genes in the identified locus, *CYP1B1* and *RMDN2*.

**ADNI DNA methylation data:** In ADNI, Illumina EPIC chips (Illumina, Inc., San Diego, CA, USA) were used to profile DNA methylation in 1,920 blood or buffy coats samples including 200 duplicate samples according to the Illumina protocols. A detailed protocol describing DNA measurement has been described previously in detail[67-69]. In brief, genomic DNA samples were obtained from NCRAD (National Centralized Repository for Alzheimer's Disease and Related Dementias), and normalization and quality control were conducted. One sample out of the total 1,920 was excluded because it had no CpG calls, and four samples were excluded because $\geq 1\%$ of the CpG sites had a detection p-value > 0.05. The remaining longitudinal 1,915 samples were normalized using the dasen method in the wateRmelon R package[70]. After normalization, quality control, and removal of duplicates, the dataset (N=1,708) of comprised beta-values was used for further analyses. We performed methylation quantitative trait loci (meQTLs) of rs2113389 with CpGs in *CYP1B1* at the first visit (N=634) using multivariate linear models adjusted for age, sex, cell composition changes, and DNA storage/source.

**AD pathology mouse model analysis:** hTau mouse model: The generation of hTAU mice was previously described[71,72]. The hTAU mice generated by Andorfer et al., (Jackson Laboratory #005491)[73] was crossed to *Mapt* knockout mouse (Jackson Laboratory #007251). The mice were bred and housed in specific-pathogen-free conditions. Both male and female mice were used. Animals used in the study were housed in the Stark Neurosciences Research Institute Laboratory Animal Resource Center, Indiana University School of Medicine. All animals were maintained, and experiments were performed in accordance with the recommendations in the Guide for the Care and Use of Laboratory Animals of the National Institutes of Health. The protocol was approved by the Institutional Animal Care and Use Committee (IACUC) at Indiana University School of Medicine. Brain extraction and tissue processing: Mice were euthanized by perfusion with ice-cold phosphate-buffered saline (PBS) following full anesthetization[74]. Brains were immediately removed. The left hemisphere was dissected into cortex and hippocampus and immediately frozen in dry ice. Samples were stored at -80°C until further processing for biochemical experiments. The frozen brain tissue was homogenized in buffer consisting of 20 mM Tris-HCl (pH=7.4), 250 mM sucrose, 0.5 mM ethylene glycol-bis (β-aminoethyl ether)-N,N,N',N'-tetraacetic acid (EGTA), 0.5 mM ethylenediaminetetraacetic acid (EDTA), and RNase-free water and were stored in an equal volume of RNA STAT-60 (Amsbio) at -80°C until RNA extraction was performed. RNA was isolated by chloroform extraction and purified using the Purelink RNA Mini Kit (Life Technologies)[75]. The cDNA was prepared from 750 ng of RNA using the High-Capacity of RNA-to-cDNA kit (Applied Biosystems), and qPCR was performed on the StepOne Plus Real-Time PCR system (Life Technologies) with the Taqman Gene Expression Assay (Cyp1b1; Mm00487229_m1, Life Technologies). Relative gene expression was determined with the △△CT method and was assessed relative to GAPDH (Mm99999915_g1). Student's t test was performed for qPCR results comparing C57BL/6J (B6; wild type) and hTAU mice. rTg4510 and J20 mouse model: Transgenic mice harboring human tau (rTg4510) and amyloid precursor protein (J20) mutations were used to investigate if gene expression changes of *Cyp1b1* was associated with the progression of Alzheimer's disease (AD) pathology[76]. The rTg4510 mouse model overexpresses a human mutant (P301L) form of the microtubule-associated protein tau (MAPT) for progressive tau pathology[77,78]. The J20 mouse line expresses a mutant (K670N/M671L and V717F) form of the human amyloid precursor protein (APP) for amyloid pathology[79,80]. The experimental models and methods were described in detail in a previous publication[76]. Briefly, all animal procedures were carried out at Eli Lilly and Company. Only female mice were used in this study[81,82]. rTg4510 (rTg(tet-o-TauP301L)4510) were bred on a mixed 129S6/SvEvTac + FVB/NCrl background[77,78]. Bi-transgenic female mice and littermate controls (WT) at ages 2, 4, 6 and 8 months-old (n = 9-10 animals per group) were used for this study. J20 (B6.Cg-Zbtb20Tg(PDGFB-APPSwInd) 20Lms/2Mmjax) were bred on a C57BL/6JOlaHsd background[80,83]. Hemizygous females and littermate controls (WT) at ages 6, 8, 10 and 12 months of age (n = 9-10 animals per group) were used for this study. Histopathology: The right hemisphere from all animals was processed and embedded in paraffin wax. Negative and positive controls were used for each immunohistochemistry experiment. To assess tau pathology, we used mouse monoclonal PG5 as the primary antibody, which recognizes tau phosphorylated at Ser409[84,85]. We also assessed tau pathology with the mouse monoclonal AT8 primary antibody specific for tau phosphorylated at Ser202 and Thr205. To assess amyloid pathology, we used mouse monoclonal biotinylated 3D6, which binds to the amino acids 1-5 in Aβ[86]. RNA isolation and highly parallel RNA sequencing: Tissue samples from each model were processed separately and individual samples were randomized to ensure that each group was equally represented in each processing batch. Total RNA from the entorhinal cortex was isolated using the AllPrep DNA/RNA Mini Kit (QIAGEN), with minor modifications to the manufacturer's protocol. RNA quality and quantity for all samples was checked using RNA ScreenTape (Agilent). The optimal eight entorhinal cortex samples for each group were selected for transcriptional profiling (total n = 128 samples; two models (rTg4510/J20) x two groups (TG/WT) x four timepoints x eight individual animals per group). Stranded-specific mRNA sequencing libraries and cDNA libraries were prepared, and libraries were individually cleaned up[87]. Samples were pooled together to a 2nM concentration, for subsequent sequencing. Pooled libraries were quantified, and final library pools were distributed across twelve HiSeq2500 (Illumina) lanes (six lanes for each model) and subjected to 125bp paired-end sequencing yielding a mean untrimmed read depth of ~20 million reads/sample. RNA-seq data processing: Raw files were demultiplexed into FASTQ files and checked for potential contamination. The randomized FASTQ files underwent quality control (QC) assessments using *FastQC*[88]. Trimming (ribosomal sequences removal, quality threshold

20, minimum sequence length 35) was performed with *fastqmcf* and trimmed samples were aligned to the *mm10* (GRCm38.p4) reference mouse genome using *STAR*[89]. Gene expression quantification was achieved using *featureCounts*[90]. RNA-seq gene expression analysis: Read counts were analyzed for differential expression using the R package DESeq2[91]. We were interested in detecting both genotype effects and progressive changes across age between the transgenic and wild-type samples. We used the following statistical model, including main effects for both Genotype and Age (both coded as categorical variables) and an interaction between these two terms:

Gene expression = Genotype + Age + Genotype*Age

To identify significant genotype effects, a Wald test was used, and to identify significant effects of age and interaction effects (i.e., Genotype*Age), we used the likelihood-ratio test, both applied with the DESeq function from the DESeq2 package[91]. To test for associations between gene expression and measures of neuropathology quantified using immunohistochemistry, we fitted linear models using DESeq and used a Wald test to calculate *P* values. These models were fitted separately for neuropathology data measured in the entorhinal cortex for each individual. *P* values were adjusted for multiple testing using the false discovery rate (FDR) method[52].

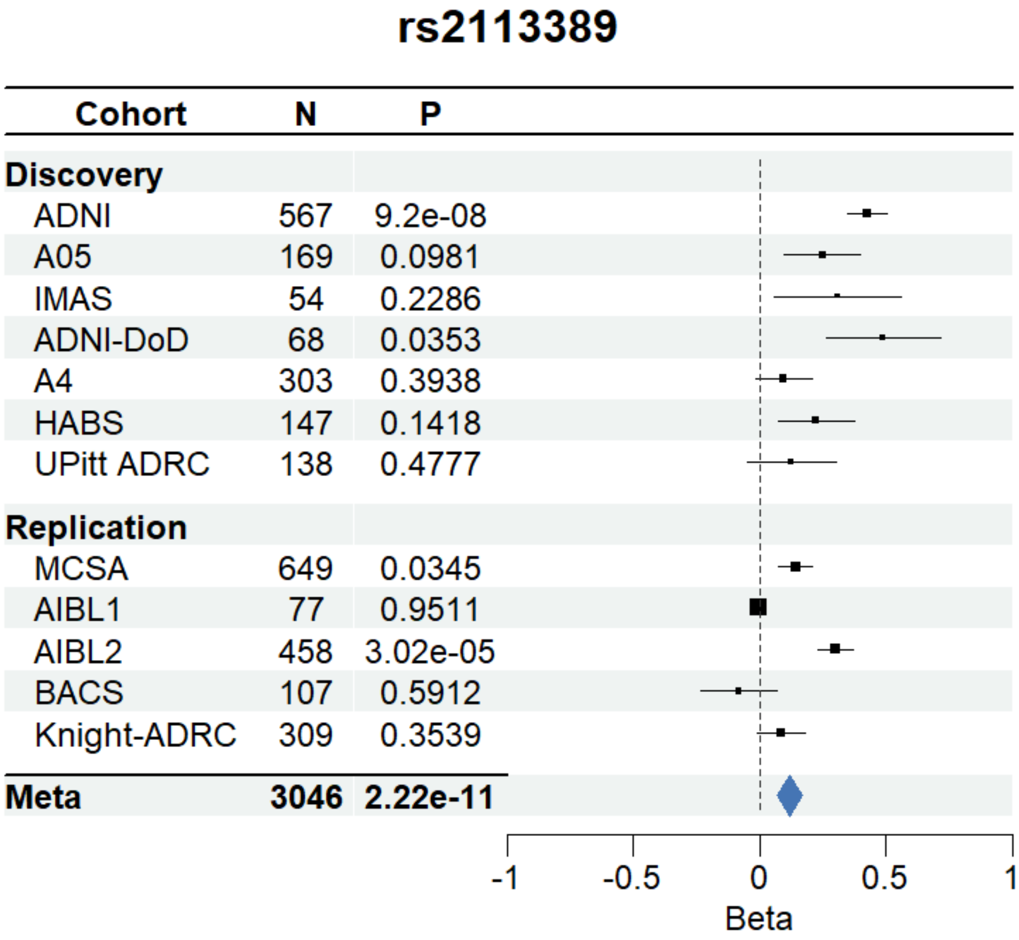


**Supplemental Figure 1.** Forest plot of rs2113389 across all included cohorts

In the discovery sample (ADNI, A05, IMAS, ADNI-DoD, HABS, UPitt ADRC), the minor allele T of rs2113389 (MAF=0.146) was associated with higher tau (Z score=5.68; *p*-value=1.37 x $10^{-8}$). We conducted a replication meta-analysis in five additional cohorts (n=1,687). The top SNP (rs2113389) in the discovery stage was replicated with the same association direction (Z score=3.83, *p*-value=1.26 x $10^{-4}$). The overall meta-analysis showed a genome-wide significant association of rs2113389 with cortical tau deposition

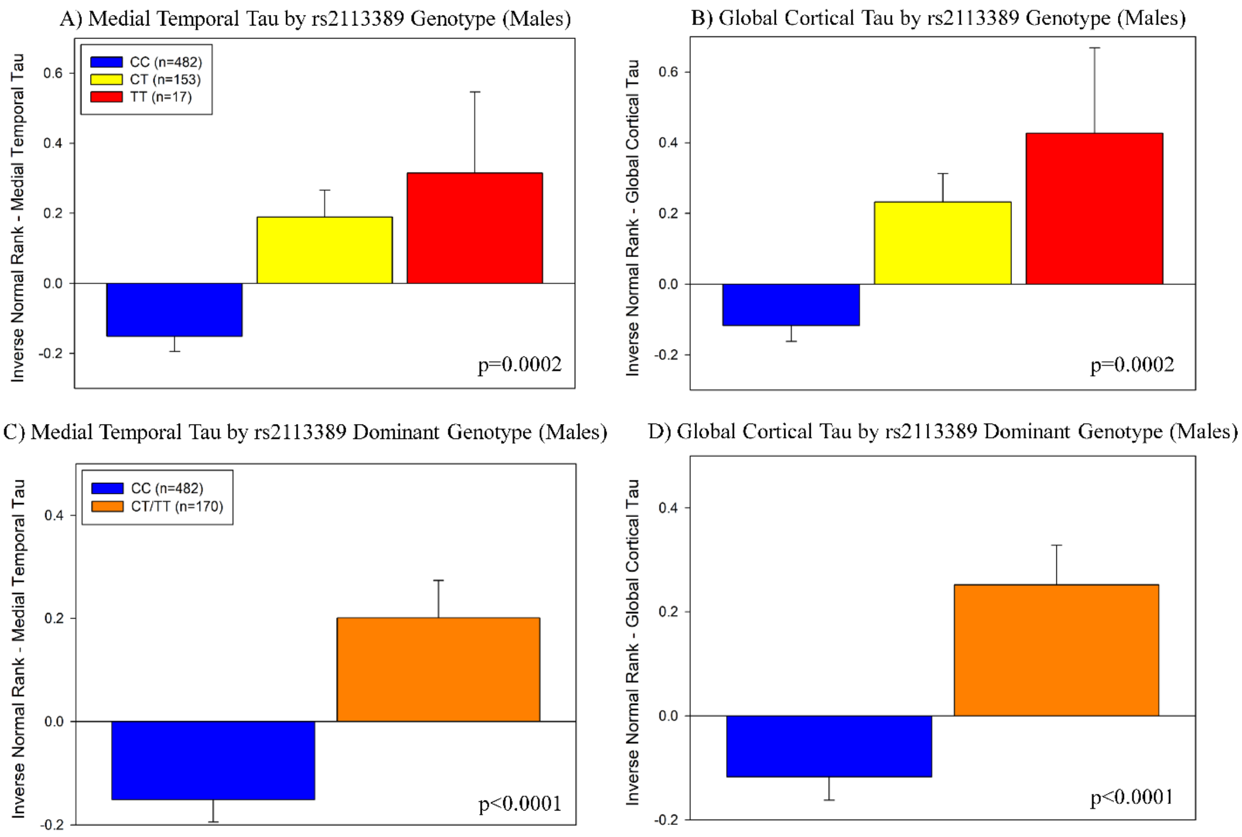


**Supplemental Figure 2.** Association of rs2113389 genotype with medial temporal and cortical tau deposition in males

In both additive and dominant models, rs2113389 genotype was associated with medial temporal lobe (MTL) and cortical tau deposition in males (additive, p=0.0002; dominant, p<0.0001). In the additive model, CT and TT individuals showed more tau deposition in the MTL (A) and cortex (B) than CC individuals, but no differences between CT and TT groups. In the dominant model, carriers of the minor allele (T) of rs2113389 have higher MTL (C) and cortical (D) tau deposition than CC individuals. *Note: tau measured as an inverse normal transformed variable of medial temporal and cortical tau SUVR*

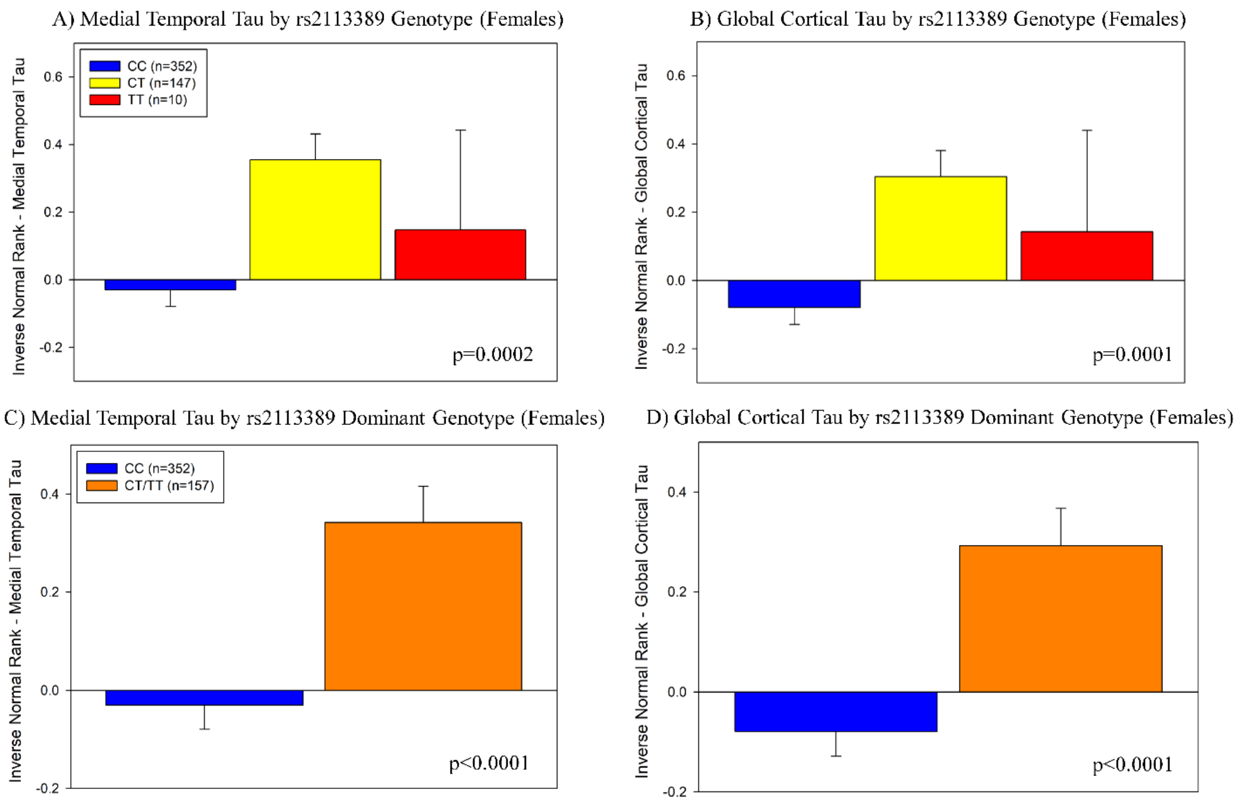


**Supplemental Figure 3.** Association of rs2113389 genotype with medial temporal and cortical tau deposition in females

In females, rs2113389 genotype is associated with medial temporal lobe (MTL) and cortical tau deposition in males (additive, $p<0.001$; dominant, $p<0.0001$). In the additive model, CT and TT individuals showed more tau deposition in the MTL (A) and cortex (B) than CC individuals, but no differences between CT and TT groups. In the dominant model, carriers of the minor allele (T) of rs2113389 have higher MTL (C) and cortical (D) tau deposition than CC individuals.

*Note: tau measured as an inverse normal transformed variable of medial temporal and cortical tau SUVR*

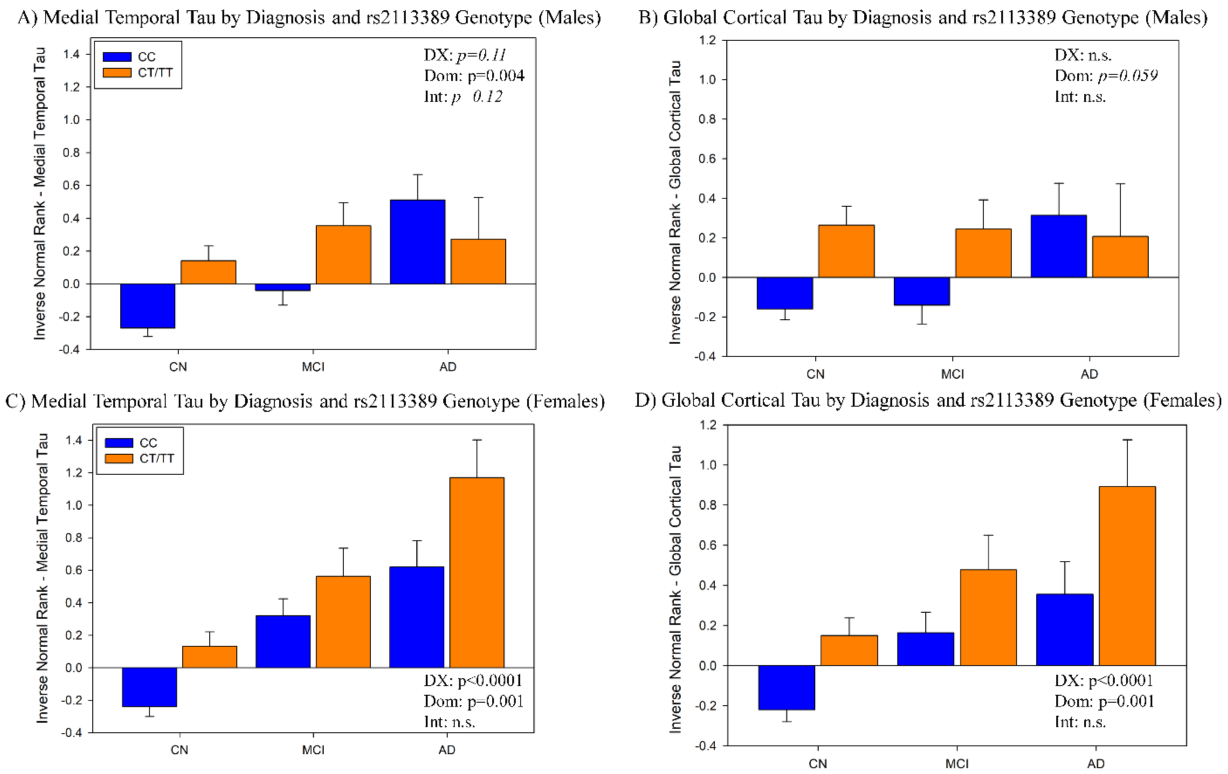


**Supplemental Figure 4.** Association of diagnosis and rs2113389 genotype with medial temporal and cortical tau deposition in sex-stratified samples

Significant main effects of diagnosis and rs2113389 dominant genotype, but no interaction effects, were observed in females (diagnosis, p<0.0001; dominant genotype, p=0.001) but not in males. Specifically, males with at least one copy of the minor allele (T) at rs2113389 showed higher tau deposition in the medial temporal lobe (MTL; A, p=0.004) and a trend for higher tau in the cortex (B; p=0.059) relative to males with a rs2113389 CC genotype. However, diagnosis was not significantly associated with MTL or cortical tau in this subsample of males only. Additionally, no interaction effects were observed in males. In females, AD participants who carried at least one minor allele (T) at rs2113389 showed the highest level of tau deposition relative to all other groups in both the MTL (C) and cortex (D). AD=Alzheimer's disease;

CN=cognitively normal; DX=diagnosis; Dom=rs2113389 dominant genotype (CC vs. CT/TT); Int.=interaction; MCI=mild cognitive impairment; n.s.=not significant; *Note: tau measured as an inverse normal transformed variable of medial temporal and cortical tau SUVR*

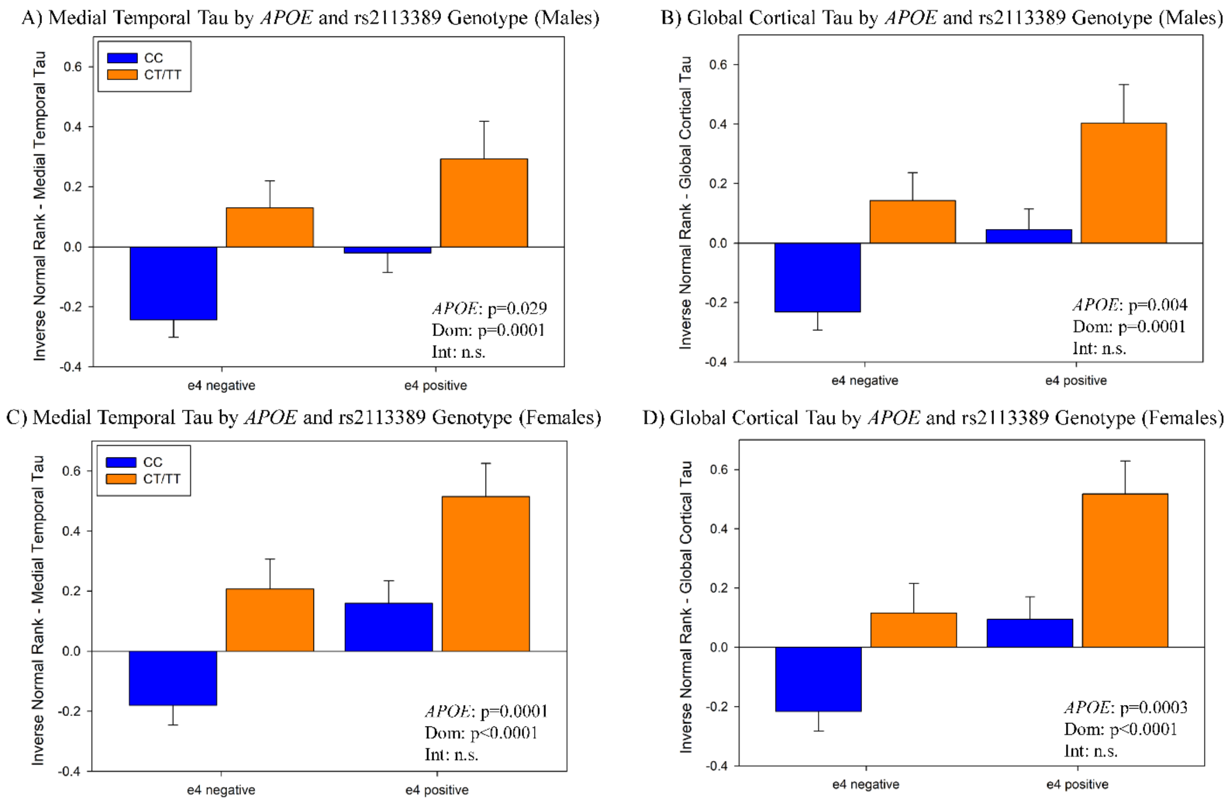


**Supplemental Figure 5.** Association of *APOE* ε4 carrier status and rs2113389 genotype with medial temporal and cortical tau deposition in sex-stratified samples

In both males and females, *APOE* ε4 carrier status and rs2113389 dominant genotype were significantly associated with medial temporal and cortical tau deposition, but no interaction effect was observed. The main effects of both *APOE* and rs2113389 genotype were stronger in female participants relative to males. Specifically, male participants who were positive for both the *APOE* ε4 allele and the minor allele (T) at rs2113389 demonstrated greater tau deposition in the medial temporal lobe (MTL; A; *APOE*, p=0.029, rs2113389, p=0.0001) and cortex (B; *APOE*, p=0.004, rs2113389, p=0.0001). Similar but even stronger results were observed in females, with the highest tau level observed in carriers of both at least one ε4 allele and minor allele (T) at rs2113389 in the MTL (C; *APOE*, p=0.0001; rs2113389, p<0.0001) and cortex (D;

*APOE*, p=0.0003; rs2113389, p<0.0001. No interactions were observed in males or females. APOE=apolipoprotein E; Dom=rs2113389 dominant genotype (CC vs. CT/TT); Int.=interaction; MCI=mild cognitive impairment; n.s.=not significant; *Note: tau measured as an inverse normal transformed variable of medial temporal and cortical tau SUVR*

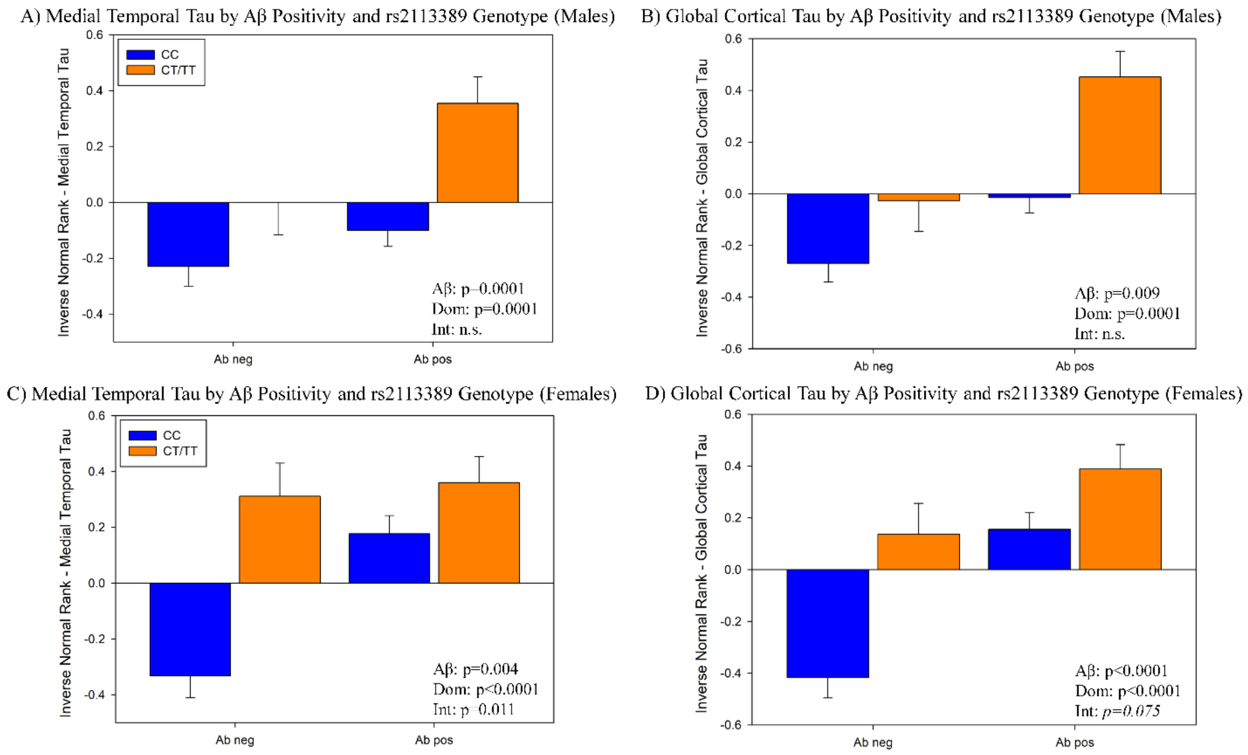


**Supplemental Figure 6.** Association of Aβ positivity and rs2113389 genotype with medial temporal and cortical tau deposition in sex-stratified samples

Males showed significant main effects of both Aβ positivity and rs2113389 dominant genotype on medial temporal (A; Aβ positivity, p=0.0001) and cortical (B; Aβ positivity, p=0.009) tau deposition are observed (rs2113389, both p=0.0001), with Aβ positive individuals carrying at least one minor allele (T) at rs2113389 showing the highest level of tau deposition relative to all other groups. No interaction effect was observed between Aβ positivity and rs2113389 in males. However, in females both main effects and an interaction effect between Aβ positivity and rs2113389 were observed. Specifically, Aβ positive females carrying at least one minor allele (T) at rs2113389 showed the highest levels of tau in the MTL (C; Aβ positivity, p=0.0001) and cortex (D; Aβ positivity, p=0.0001) relative to all other groups (rs2113389, both p<0.0001). An interaction effect was also seen, which was significant in the MTL (p=0.011) and showed trend significance in the cortex (p=0.075). Aβ=amyloid-beta; Dom=rs2113389 dominant genotype (CC vs. CT/TT); Int.=interaction; MCI=mild cognitive impairment; n.s.=not significant; *Note: tau measured as an inverse normal transformed variable of medial temporal and cortical tau SUVR*

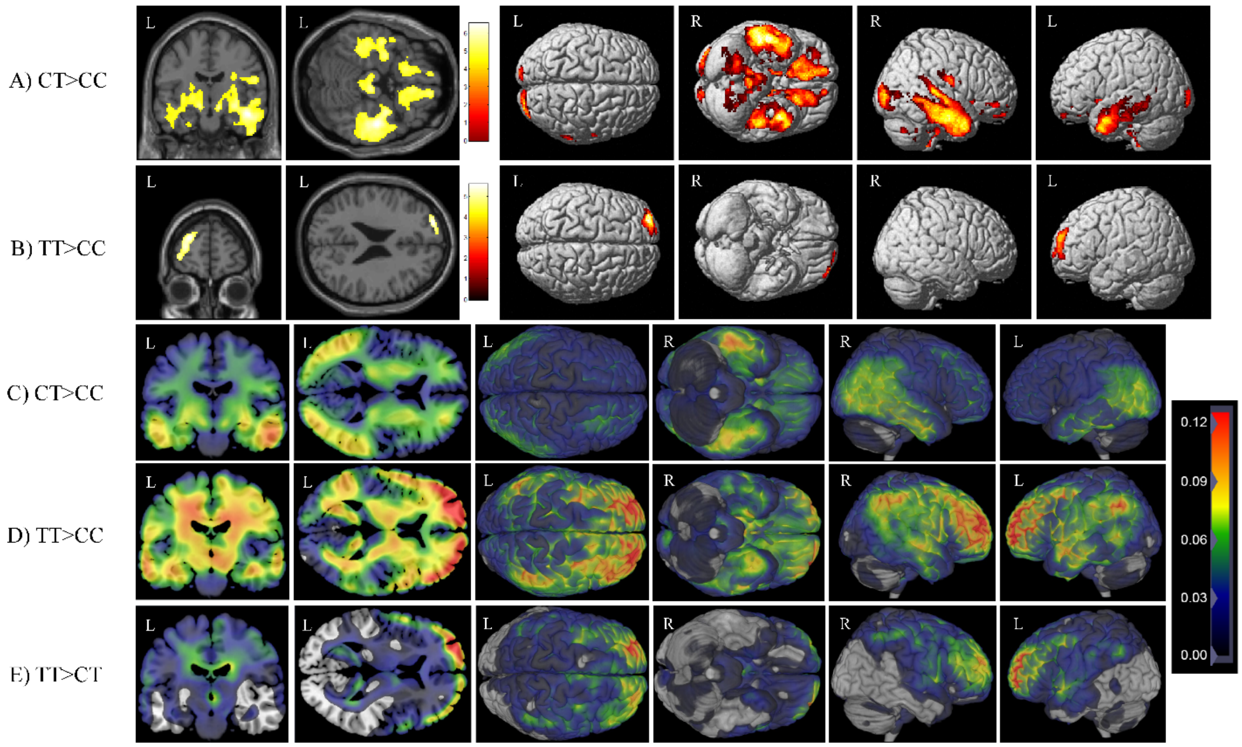


**Supplemental Figure 7.** Exploratory voxel-wise analysis of the effect of rs2113389 additive genotype on tau deposition

Using an additive model, the voxel-wise statistical maps of tau deposition showed that individuals with (A) rs2113389 CT genotype (n=300) demonstrated significantly higher tau deposition in the inferior frontal, parietal, and medial and lateral temporal lobes relative to those with a rs2113389 CC genotype (n=834). (B) Individuals with a rs2113389 TT genotype (n=27) also showed more tau deposition than those with a rs2113389 CC genotype in a focal region in the left frontal cortex. No significant differences were seen between rs2113389 CT and TT genotype individuals on voxel-wise statistical maps. The latter two findings were likely limited topographically due to the small sample size of the TT group. Images are displayed at a voxel-wise threshold of $p<0.05$ with family-wise error correction for multiple comparisons and a minimum cluster size (k)=100 voxels. Due to the very unequal group sizes, we also evaluated non-statistically limited beta-map value maps. (C) Beta-value maps suggest that rs2113389 CT heterozygotes show higher tau deposition than CC homozygotes in primarily parietal and temporal regions. (D) Alternatively, the beta-value maps suggest that rs2113389 TT homozygotes have higher tau in widespread regions of the temporal and parietal lobes, as well as strongly in the bilateral frontal lobes, relative to rs2113389 CC homozygotes. (E) Finally, beta-value maps suggested that rs2113389 TT homozygotes show greater bilateral frontal tau than rs2113389 CT heterozygotes.

A) *CYP1B1* Gene Expression B) *RMND2* Gene Expression

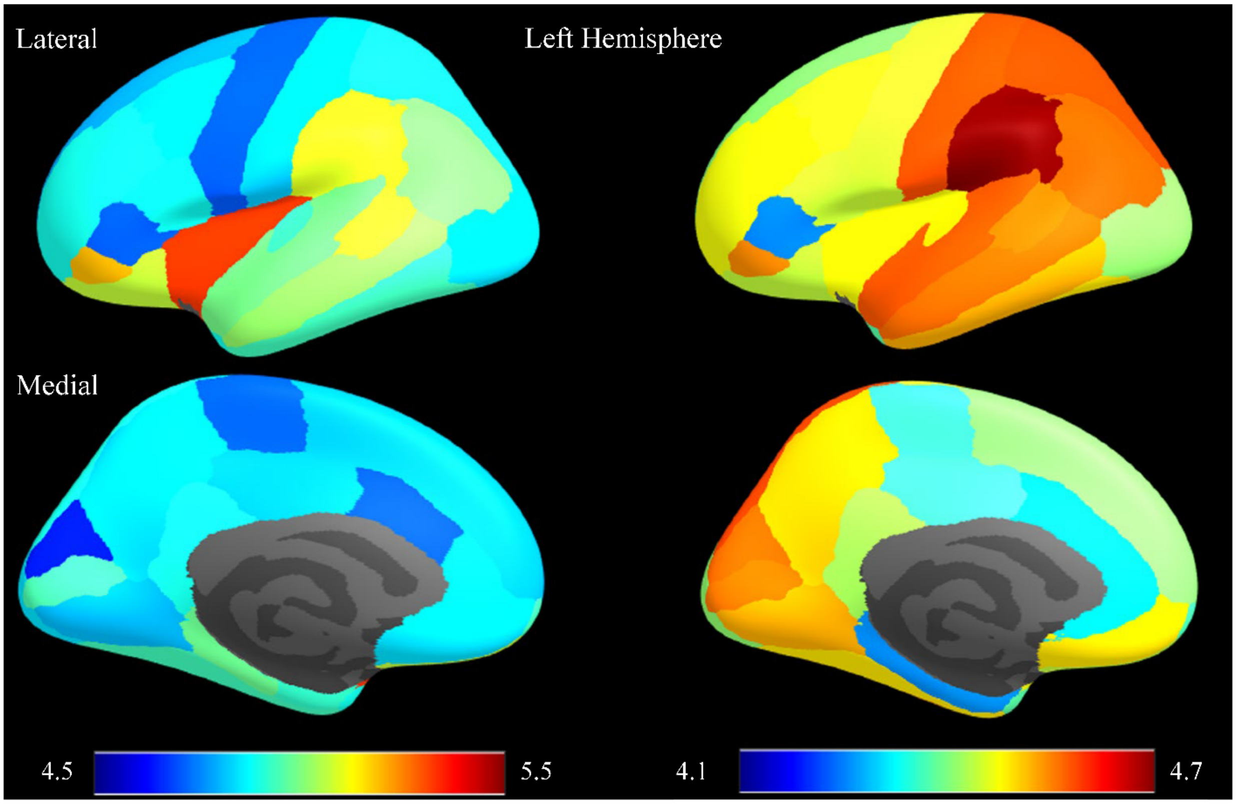


**Supplemental Figure 8.** Spatial distributions of *CYP1B1* and *RMDN2* gene expression profiles in the left hemisphere

The lateral (top) and medial (bottom) brain views for *CYP1B1* (A) and *RMDN2* (B) gene expression in the Allen Human Brain Atlas are shown. *CYP1B1* shows high gene expression in the insula, orbitofrontal cortex, temporal lobe, and medial temporal lobe. *RMDN2* shows strong gene expression levels in the temporal lobe, visual cortex, frontal (medial orbitofrontal cortex) and posterior (precuneus and isthmus cingulate cortex) default mode network regions, and sensorimotor cortex. Of note, the regions showing high gene expression levels for both genes partly overlap with the typical regions with high tau deposition in AD.
